# Are different consumer sleep technologies measuring the same essential aspects of sleep?

**DOI:** 10.64898/2026.03.31.26349815

**Authors:** Kiran K G Ravindran, Ciro della Monica, Guiseppe Atzori, Marta Messina Pineda, Ramin Nilforooshan, Hana Hassanin, Victoria Revell, Derk-Jan Dijk

## Abstract

**Study objectives:** Consumer sleep technologies (CSTs) enable low-burden longitudinal sleep monitoring, and their output measures are often interpreted as equivalent to polysomnography (PSG) measures. We applied a measurement reliability-aware approach to determine whether CST-derived ‘sleep’ measures (1) are interchangeable or device-specific, (2) can reliably assess trait-like sleep characteristics of an individual, (3) can be reduced to latent principal components of sleep, and (4) can be used for classification and biomarker discovery.

**Methods:** Data from 74 older adults (20 people living with dementia [PLWD]) were collected at-home (upto 14 nights; Total=752nights) using four tools simultaneously: research-grade actigraphy (Axivity), a wearable (Withings Watch), a nearable (Withings Sleep Analyzer) and Sleep Diary, followed by one in-lab PSG assessment. We used repeated-measures correlation analyses, intraclass correlation coefficients (ICC), principal component analysis (PCA) and binary classification models to address our objectives.

**Results:** Single-night between-device correlations and correlations with PSG were moderate (0.3≤r<0.7) for some duration- and timing-related measures, but other associations were weak (r<0.3). Seventy-one percent of sleep measures reached acceptable reliability (ICC≥0.7) within seven nights of aggregation, but the required aggregation window varied across measures, tools and between PLWD and Controls. Reliability-filtered PCA yielded stable and interpretable principal components, but Duration was the only component showing moderate between-device association. Principal components were successfully used to classify PLWD vs Controls but feature importance varied across devices.

**Conclusions:** Aggregation of CST derived measures across 7-14 nights, yielded reliable measures, most of which were device-specific, with duration being the only essential aspect transferable between devices.

**Statement of Significance:** Consumer sleep technologies (CSTs) are increasingly used to investigate relationships between sleep and brain-body health but the extent to which their measures are reliable and comparable across devices remains unclear. Using at-home data collected simultaneously from wristworn Axivity, two CSTs (a wearable and a nearable) and Sleep Diary, in older adults and people living with dementia, we show that while several duration- and timing-related sleep measures are moderately associated across devices, most other associations are weak. Approaches such as multi-night aggregation or principal component analysis improved reliability of CST-derived measures, although most measures remained device-specific. The findings imply that except for duration measures, sleep measures collected on different devices are not equivalent and care should be taken when comparing studies using different CSTs.

## Introduction

Sleep is increasingly recognised as a behavioural and physiological marker with relevance for understanding disease risk, monitoring disease progression, biomarker development and evaluating interventions for a range of health conditions across the life span, including older adults and people living with dementia (PLWD) [1, 2, 3, 4]. Investigating these associations longitudinally requires reliable sleep measurements. To facilitate continuous monitoring of sleep in naturalistic living conditions, consumer sleep technologies (CSTs) are increasingly adopted [5, 6, 7].

Most CSTs (both non-EEG wearables and nearables) estimate sleep using actigraphy or movement combined with peripheral physiological signals rather than through direct measurement of brain activity [8]. Wearables, such as wrist-worn devices, primarily rely on actigraphy combined with photoplethysmography to quantify heart and breathing rate and oxygen saturation. Nearables, such as under-mattress sensors or radar-based devices, utilise ballistography to derive activity and physiological signals such as respiration and heart rate. A key advantage of nearables is that they can collect data whether the participant is in bed or out of bed. Unlike wearables, nearables are embedded in the living environment, most commonly the bedroom, operate in a contactless manner, and require no active user engagement, making them effectively burden-free and particularly suitable for older adults and cognitively impaired populations.

The widespread availability, ease of use, relatively low cost, and digital infrastructure (automated data acquisition, cloud-based storage, and remote access) of CSTs have facilitated their rapid deployment in large observational studies, allowing continuous monitoring outside the laboratory [6, 9, 10]. These features make CSTs particularly attractive in populations where repeated sleep laboratory assessments are neither feasible nor tolerated and research approaches in which capturing habitual sleep patterns and night-to-night variability is more informative.

Most CSTs are not regulated and rely on proprietary, non-transparent algorithms. As a result, their outputs are not held to clinical performance or reporting standards. Their outputs are often labelled using terminology borrowed from polysomnography (PSG), despite fundamental differences in sensing and processing [11, 12, 13]. The use of familiar PSG labels and vocabulary can therefore create a false impression of validity and equivalence, leading to an inflated perception of the clinical importance of these measures. Consequently, similarly named sleep measures (e.g., total sleep time [TST] rapid eye movement [REM] sleep) may exhibit systematic bias, limited agreement with gold-standard assessments (i.e., PSG), and poor interchangeability across devices [14]. Without comparability across devices, identically labelled sleep measures may represent different underlying physiological or behavioural signals, reducing interpretability, preventing meaningful comparison across studies, and limiting their usefulness in clinical applications. This heterogeneity in sleep measures across devices, firmware versions, and analytical approaches represents a key limitation that challenges the development and interpretation of CST-derived digital biomarkers. Expert recommendations therefore emphasise the need to move beyond isolated, label-based sleep variables toward more harmonised, multidimensional characterisation of sleep and circadian function [15].

In general, across the numerous validation studies comparing different CSTs with PSG, CSTs tend to perform moderately for sleep timing and duration measures, while measures that depend on accurately distinguishing quiet wake from sleep (e.g., wake after sleep onset, WASO) or identifying deep sleep and REM sleep show limited agreement and larger bias [16, 17, 18, 19, 20]. These studies are predominantly conducted in younger healthy adults with relatively high sleep efficiency and fewer nocturnal awakenings, which may lead to overestimation of device performance and limit generalisability to older adults and other clinical populations with more fragmented sleep, and often do not follow standardised evaluation approaches [21].

Although rigorous validation of consumer sleep technologies against PSG is useful for understanding their measurement accuracy and for the clinical interpretation of the sleep measures, such validations are not feasible across the full range of populations, comorbid conditions, and real-world contexts. This creates a gap between what can be measured at scale and traditional validation approaches, particularly for applications that rely on longitudinal, trait-like stability of sleep measures rather than absolute agreement against gold-standard PSG, the reliability of which has been rarely assessed in specific populations such as PLWD. In this context, a measure selected as an indicator of an individual’s sleep characteristics or a sleep biomarker should provide a reliable estimate of that individual’s sleep and meaningful changes therein, rather than variability driven by measurement noise, algorithmic instability or variability inherent to CSTs [22].

There is therefore a need for a device-agnostic framework that moves beyond nominal sleep measures and focuses instead on identifying stable, and interpretable aspects of sleep that are robust to device-specific characteristics, comparable/transferable across devices and capable of quantifying an individual’s sleep characteristics. Our work presents a data-driven approach to quantify the reliability (i.e., night-to-night stability rather than absolute agreement with PSG) of automatically generated sleep measures, examine their latent structure, and evaluate their predictive utility for distinguishing PLWD from cognitively healthy older adults. Importantly, reliability can be thought of in real-world device deployment terms as the number of nights of data required to obtain a stable estimate of an individual’s sleep characteristics of interest in a longitudinal study. To provide context for interpreting CST-derived outputs, we also examine their agreement with PSG and the associations between similarly named sleep measures across devices. Using at-home data collected simultaneously from four sleep assessment modalities (actigraphy [Axivity], two Withings CSTs: a wearable and a nearable and Sleep Diary), we specifically address the following questions: (1) do identically labelled CST-derived sleep measures measure the same underlying sleep aspect, (2) which CST derived sleep measures are reliable/stable within a device and how many nights of data are required to obtain reliable estimates of an individual’s sleep characteristics, (3) whether the reliable measures converge into interpretable sleep aspects such as Duration or Continuity, (4) whether these aspects are consistent and interchangeable between devices, and (5) as an example, whether reliability relates to the predictive utility of distinguishing PLWD from cognitively intact older adults.

## Methods

### Population characteristics

A total of 74 older adults (age, mean ± SD = 70.5 ± 6.5 years) each contributed 7 to 14 nights of data at home, followed by laboratory PSG acquired at the UK Dementia Research Institute Clinical Research Facility at the Surrey Sleep Research Centre. The data were drawn from two different studies conducted at different time periods using similar methodology. Both studies received a favourable ethical opinion and were conducted in line with the Declaration of Helsinki and Good Clinical Practice (GCP). Study 1 (University of Surrey ethics: UEC-2019-065-FHMS) involved cognitively intact older adults, while Study 2 (NHS ethics: 22/LO/0694) also involved people living with dementia (PLWD), their study partners and cognitively intact older adults. Both cognitively intact older adults and study partners of PLWD were considered as Controls. The PLWD were diagnosed based on prodromal or mild Alzheimer’s disease cognitive test results and CT/MRI imaging. A detailed description of the protocols and inclusion/exclusion criteria can be found in [23]. Some aspects of these studies have been reported previously [6, 24, 25].

### Sleep assessment modalities

In both studies, participants kept a consensus Sleep Diary (SD) and wore a research-grade accelerometer (Axivity Ltd, AX; model AX3/6) alongside two Withings (Withings, France) CSTs: the Withings Sleep Analyser (WSA) and the Withings Watch (WW). The WSA is an under-mattress pneumatic nearable that uses ballistographic sensing, while the WW is a wrist-worn wearable (smartwatch) that combines activity tracking and photoplethysmography, similar to other wearable CSTs. Although sleep diaries are known to be susceptible to recall bias, particularly in older adults and PLWD, we have included it in our analyses to provide a comparison with CSTs as it is a widely used method of subjective sleep assessment [26].

The AX data were collected at 100 Hz to allow up to 14 nights of continuous at-home monitoring, and sleep/wake labels were generated using the Biobank accelerometer analysis package to estimate the sleep measures [27]. The Axivity was selected because it is a widely used research-grade accelerometer, including in the UK Biobank cohort. A Sleep Diary–free approach was adopted for AX-derived sleep estimation, as the use of sleep diaries is not always feasible in all populations. Sleep periods were identified within each noon-to-noon window, and periods separated by wake periods of less than 60 minutes were merged to account for nocturnal awakenings. The merged sleep episode with the longest duration within each noon-to-noon window was defined as the diary-free sleep period time (SPT). The start and end times of this episode correspond to sleep onset and final awakening, respectively. Sleep periods outside this window were treated as naps.

The WSA and WW automatically generated their proprietary sleep summary measures for every in-bed or recording period detected. The data were downloaded using the Withings application programming interface (API) as JSON files. The data collected from the four tools/devices were extracted and curated into a tabular format using MATLAB. All available sleep measures provided by each device were used in the analysis. Measures common across the devices included total sleep time (TST), sleep period time (SPT), sleep onset latency (SOL), sleep efficiency (SEFF), wake after sleep onset (WASO), number of awakenings (NAW), and sleep timing measures (for example, lights off/on or sleep onset and final awakening). All subsequent analyses were performed using Python (v3.11) and its associated data analysis libraries. The sleep measures corresponding to the four devices are denoted by the following prefixes: s – Sleep Diary; x – Axivity; m – Withings Sleep Analyser; w – Withings Watch. The complete list of all the measures used is listed in the Supplemental Materials, Table S1.

### Measurement framework for estimating the latent sleep state

Sleep is a latent neurophysiological state of the brain and body that cannot be observed directly (Figure 1A). In clinical sleep research, PSG provides the reference gold standard by recording both central nervous system, i.e., electroencephalogram, and peripheral physiological signals, such as muscle tone, that are inspected visually by a trained human scorer and translated into discrete 30-second sleep stage labels (Wake, N1, N2, N3 and rapid eye movement sleep [REM], with N1, N2 and N3 collectively representing non-rapid eye movement sleep [NREM]) using standardised (American Academy of Sleep Medicine, AASM) scoring rules and then summarised into sleep measures such as TST, WASO and N3 duration. Thus, PSG-derived sleep measures are themselves estimates of the underlying sleep/vigilance states rather than direct measurements of latent “true” sleep/vigilance states. In the PSG sleep estimation process, systematic errors arise from the discretisation of the continuous sleep physiology based on standard rules over fixed 30-second epochs, while random errors arise from scorer variability, artefacts and the presence of ambiguous epochs around sleep stage transitions.

**Figure 1.**
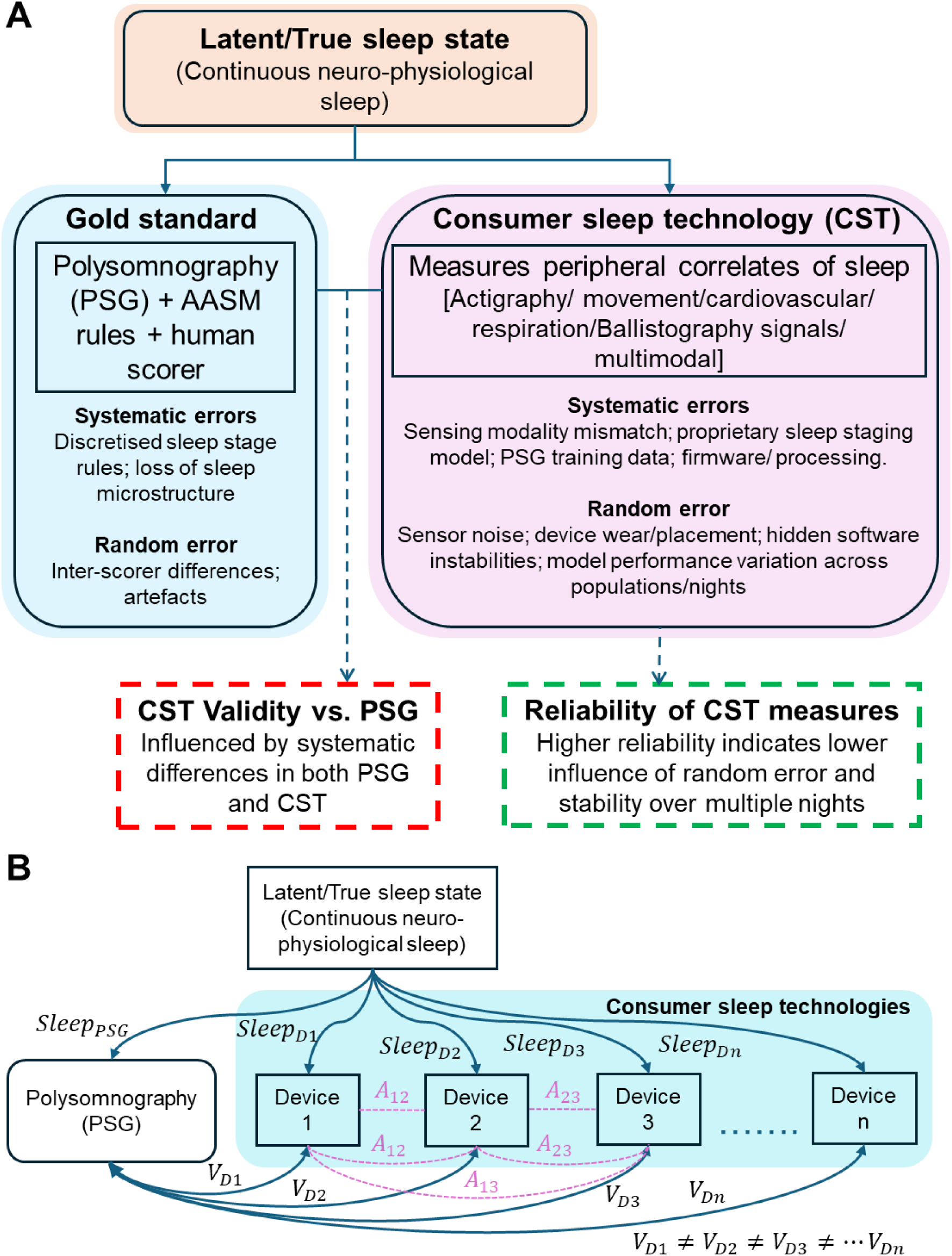
Measurement framework for estimating the latent sleep state. **A.** Polysomnography (PSG) and consumer sleep technologies (CSTs) measurement pathways and sources of error. **B.** Illustration of device-specific proprietary representation of latent sleep and corresponding errors leading to differing validity of CSTs against PSG. 𝑆𝑙𝑒𝑒𝑝_𝑥_ depicts the representation of latent sleep captured by PSG or Device (D), 𝑉_𝐷𝑥_ depicts the validity of a device against PSG and 𝐴_𝑥𝑥_ depicts the association between devices.

CSTs, on the other hand, estimate sleep using peripheral physiological and behavioural signals, moving further away from latent sleep state features that originate from the brain. Rather than directly measuring the neurophysiological signals which are essential to define conventional sleep stages, CSTs determine sleep states from signals such as movement, cardiovascular, respiration, or other modality-specific correlates. These signals are then further processed through device-specific hardware, firmware, and proprietary sleep estimation algorithms generally trained using PSG-derived discrete sleep stage labels.

Further, since different CSTs use different sensing modalities and proprietary signal processing and sleep estimation pathways, they cannot be assumed to measure identical representations of sleep. Instead, each device extracts its own distinct set of peripheral physiological correlates and a device-specific representation of latent sleep (𝑆𝑙𝑒𝑒𝑝_𝐷𝑥_). Thus, differences in validity of a CST device against PSG (𝑉_𝐷𝑥_) may reflect not only differences in device performance, but also differences in the aspects of peripheral physiology captured by each device, the systematic errors introduced by the unique sensing modality and PSG-dependent sleep staging models and the mismatch between device-specific and PSG-derived sleep representations. Pairwise associations between CSTs (𝐴_𝑥𝑥_) indicate the extent to which different device-specific representations of sleep compared to each other, rather than whether they measure the same latent sleep state in the same way (Figure 1B).

Within this measurement framework, random error in CST-derived sleep measures may arise from sensor noise, device wear or placement, environmental influences, firmware/ software changes over the measurement period and night to night instability and population-specific performance differences due to the training data used. As the systematic error or bias in sleep measurement is inherent to each device-specific representation of sleep, it cannot be fully removed or adjusted for. In this context, reliability becomes an important property of a CST-derived sleep measure, indicating whether it provides sufficiently stable estimates for longitudinal monitoring and development of digital biomarkers at the individual level. For this reason, the present study focused on characterising the reliability of sleep measures across devices and sleep domains.

### Laboratory validation against PSG and at-home between-device associations

The SomnoHD system (SOMNOmedics GmbH^TM^, Germany) was used to collect the PSG data during the laboratory session. Sleep staging was performed as per the (AASM) guidelines, and a consensus hypnogram was generated by two scorers (a Registered Polysomnographic Technologist™ [RPSGT] and a trained independent scorer) [28]. The sleep measures were estimated using the consensus hypnogram, and the association between the PSG and the Sleep Diary, Axivity, and Withings-derived sleep measures was examined using Bland-Altman agreement measures, as described previously in [16] and the Supplemental Materials. In addition, Pearson’s correlations (weak: 0.10 to 0.29; moderate: 0.30 to 0.7; and strong: > 0.7) were computed to examine the associations between devices in the laboratory. The validation of a portion of WSA sleep measures (35 participants) has already been published in [16] and is reused here.

To provide additional context for interpreting similarly labelled sleep measures across devices, we explored associations between sleep measures at-home using a repeated-measures correlation approach (*rm_corr* function in the pingouin package, Python), which accounts for the fact that multiple nights of data were collected from the same participants. These associations were computed between unique pairs of sleep measures within the same device and across devices. Rather than correlating values across individuals, this method estimates whether night-to-night changes within an individual in one measure are accompanied by corresponding changes in another measure. Because a large number (56 measures across four devices) of pairwise correlations were evaluated, multiple testing correction was performed using the Benjamini-Hochberg false discovery rate (FDR) procedure, with adjusted q-values used to assess statistical significance (q < 0.05). Repeated measures correlations were interpreted using the following thresholds: weak: 0.10 to 0.29; moderate: 0.30 to 0.7; and strong: > 0.7.

### Reliability analysis of sleep measures

The reliability of the sleep measures was assessed using intraclass correlation coefficient, ICC(1,1) (one-way random-effects, single-measure), which reflects the reliability of a single night of measurement. ICC(1,1) quantifies the proportion of total variance attributable to stable between-participant differences across nights. All available nights of data were used in the estimation of the variance components, but the resulting ICC(1,1) represents the reliability expected from a single-night estimate of a sleep measure from a given device. The ICC was calculated separately for each device and sleep measure. It is defined as,

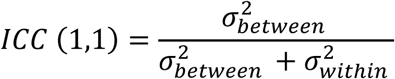

Here, *σ_between_*^2^ denotes between-participant variance and *σ_within_*^2^ denotes within-participant variance [29]. In this framework, non-systematic measurement error is reflected in the within-participant variance component and represents variability that is not attributable to stable inter-individual differences.

Because such variability increases the within-participant variance and consequently lowers the ICC estimates, we evaluated the effect of multi-night (k) aggregation on reliability (ICC(k,1)) using a Spearman-Brown prediction formula [29, 30, 31]. This approach estimates expected reliability under repeated measurements without recomputing ICCs from aggregated data. The modified Spearman-Brown formula for estimating the number of nights (k) required to achieve a target reliability (𝐼𝐶𝐶_𝑇_) given single-night reliability (ICC(1,1) or ICC) is defined as,

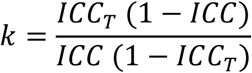

When within-participant variance is predominantly driven by physiological night-to-night variability, as is expected for continuity measures such as WASO, averaging over multiple nights is expected to improve ICC estimates. In contrast, measures dominated by device-related measurement noise or algorithm-related noise are expected to show little or no improvement with aggregation [32]. Accordingly, reliability was evaluated across increasing numbers of nights (up to 14) using Spearman-Brown extrapolation to assess whether aggregation improved measurement stability [33]. Measures that failed to achieve acceptable reliability (ICC ≥ 0.7) after aggregation were considered unlikely to reflect stable or interpretable sleep characteristics. The use of an ICC threshold of 0.7 here is intended to illustrate the proposed framework rather than to define a general threshold [33, 34].

In addition to ICC, we quantified the ratio of within to between-participant variance to better understand the stability of the measures. Higher ratios indicate greater night to night variability relative to between-participant differences and thus poor reliability. We also computed the change in ICC between single-night and seven-night estimates (ΔICC) to assess how much reliability improves with aggregation.

Associations between reliability metrics across all sleep measures were assessed using Spearman rank correlations, examining relationships between single-night ICC, within-to-between participant variance ratio, and the number of nights required to reach ICC ≥ 0.7. Because reliability metrics were non-normally distributed and heteroscedastic, group comparisons were performed using Mann-Whitney U tests with Cliff’s delta as a non-parametric effect size [35]. To assess whether reliability characteristics differed systematically across devices and participant groups, measure-level reliability metrics were modelled using linear mixed-effects models, with device and participant group included as fixed effects and sleep measure included as a random intercept.

### Identifying interpretable sleep aspects

To identify latent and interpretable sleep aspects across devices, we applied principal component analysis (PCA) to the sleep summary measures followed by orthogonal Varimax rotation to simplify the component structure and allow better interpretability. This approach is consistent with previous work on the application of Varimax-rotated PCA to longitudinal sleep measures [2, 36]. PCA was performed on both device-specific and pooled device data and components with eigenvalues greater than one were retained following visual inspection of the scree plots.

The data were standardised (z-scored) before PCA. Components were interpreted as latent sleep aspects based on the pattern and magnitude of their loadings, with absolute loadings greater than 0.45 considered meaningful contributors. This approach was used to identify interpretable and consistent patterns within the sleep measures provided by each device. These components were labelled to reflect dominant sleep domains such as sleep timing, duration, continuity and napping. To assess the temporal stability of these derived sleep aspects, component scores were computed at the night level and their reliability was quantified using ICC. This enabled comparison between the stability of individual sleep measures and that of the derived latent sleep aspects. Here, ICC was interpreted as an index of within versus between-participant stability of the latent sleep aspects, rather than as a measure of device-level reliability. This approach allowed evaluation of whether combining multiple sleep variables into latent dimensions improved interpretability and stability relative to individual sleep measures.

### Association between PCA-derived sleep aspects

To examine whether the derived sleep aspects were shared across devices, we explored associations between PCA-derived sleep aspects using a repeated-measures correlation approach. These associations were computed between unique pairs of PCA-derived sleep aspects across devices. This approach was applied consistently to both raw and clean (reliability filtered, i.e., measures that failed to achieve acceptable reliability [ICC ≥ 0.7] were removed) PCA-derived sleep aspects, allowing direct comparison of associations before and after reliability filtering. By modelling within-participant variation, the analysis avoids inflation of correlations that can arise when individuals differ systematically in their overall sleep levels. Multiple testing correction was performed as explained in the section, Laboratory validation against PSG and at-home between-device associations.

### Predictive utility of the measures

The predictive utility of CST-derived sleep measures was evaluated using binary classification models based on extreme gradient boosting (XGBoost) to distinguish PLWD from Controls. Models were constructed using both raw sleep measures and PCA-derived sleep aspects, with analyses repeated separately for each device. Model performance was evaluated using five-fold cross-validation with participant-level splitting to ensure that all observations from a given participant were confined to either the training or test set. Performance was summarised using the area under the receiver operating characteristic curve (AUC), which reflects how well the model discriminates between Controls and PLWD and average precision (AP), which reflects how accurately the model identifies true PLWD labels [37].

To aid interpretation of model behaviour, SHapley Additive exPlanations (SHAP) were used to quantify the contribution of individual sleep measures and sleep aspects to model predictions. This allowed identification of features that consistently contributed to classification performance across cross-validation folds. Finally, SHAP-derived feature importance was compared with reliability metrics, including intraclass correlation coefficients and within-to-between variance ratios, to examine whether sleep measures and sleep aspects with higher measurement reliability contributed more strongly to predictive performance.

## Results

### Population and data characteristics

Seventy-four participants (mean age ± SD = 70.5 ± 6.5 years; 31 [42%] women) contributed to the at-home or in-lab data. The analysis data comprised 54 cognitively intact controls (mean age= 69.9 ± 6.7 years; 25 [46%] women) and 20 PLWD (mean age= 72.2 ± 5.7 years; 6 [30%] women). The detailed demographic characteristics of the study population are presented in Table 1. Based on self-report measures, the majority of participants did not report significant sleep disturbance (72%, n = 53; PSQI ≤ 5), and only a small proportion reported excessive daytime sleepiness (4%, n = 3; ESS > 10). According to PSG, nearly half of the participants (35/74, 47%) had moderate-to-severe apnoea (apnoea-hypopnea index (AHI) ≥ 15).

**Table 1.** Population Characteristics. Values shown are mean followed by (standard deviation) and [min, max].

| **Characteristics** | **Total** | **PLWD** | **Controls** |
| --- | --- | --- | --- |
| **N** | 74 | 20 | 54 |
| **Age (years)** | 70.5 (6.5)<br>[44 83] | 72.2 (5.7)<br>[61 81] | 69.9 (6.7)<br>[44 83] |
| **Women (n [%])** | 31 [42] | 6 [30] | 25 [46] |
| **Standardized Mini-Mental State Examination (SMMSE)** | 28.3 (1.5)<br>[24, 30] | 27.1 (1.6)<br>[24 30] | 28.7 (1.2)<br>[25 30] |
| **BMI (kg/m<sup>2</sup>)** | 26.5 (5.2)<br>[16.8 42.1] | 26.1 (6.8)<br>[16.8 42.1] | 26.7 (4.5)<br>[20 39.8] |
| **AHI (events/hr)** | 18.2 (14.4)<br>[1.6 66.7] | 20.2 (14.7)<br>[1.7 48.6] | 17.5 (14.2)<br>[1.6 66.7] |
| **Pittsburgh Sleep Quality Index (PSQI)** | 4.3 (2.9)<br>[0 14] | 4.4 (3.8)<br>[0 14] | 4.2 (2.5)<br>[0 12] |
| **Epworth Sleepiness Scale (ESS)** | 4.1 (3)<br>[0 12] | 4 (3.1)<br>[0 10] | 4.2 (3.0)<br>[0 12] |

For the at-home analysis only 65 participants with data from all four devices were available. In these 65 participants, 45 nights of data at home were missing from Axivity for the 65 participants resulting in a total of 752 nights (mean ± SD = 11.6 ± 2.8 nights) included in the analysis. The definitions of Withings sleep measures are available in the Withings API reference, and the complete list of sleep measures and definitions are provided in Supplemental Table S1. There were 56 sleep measures, including 15 from Sleep Diary, 11 from Axivity, 18 from Withings Sleep Analyser and 12 from Withings Watch (Supplemental Materials, Table S1). The sleep measures that had more than 2% missing data were WW Sleep_Score (23.9%), Sleep Diary Rest upon awakening (sRuA, 6.5%), WSA AHI (6.3%) and Sleep Diary SOL (2.7%).

An example data snapshot collected over 14 nights at home is visualised in Figure 2. In Figure 2A, using data from all devices (with Sleep Diary information shown as inverted triangles and a solid navy-blue line), it can be seen that the participant had a consistent bedtime and this is accurately captured by the contactless under-mattress device (WSA), while the wearable accelerometer (Axivity) and smartwatch (WW) show larger differences in their recording times. TST varied between the devices (Figure 2B and 2C). The self-reported TST (mean±SD= 520.4 ± 38.8 minutes) was the highest, followed by WSA (513.9 ± 31.7 minutes), WW (510.7 ± 67.9 minutes) and Axivity (442.3 ± 54.8 minutes). In this participant, it is noteworthy that the direction of the night-to-night variation can differ between devices, with each device ranking a different night as the longest. For example, Sleep Diary and WSA ranked Day -2 as the longest, while for Axivity it was Day -14, and for WW it was Day -12.

**Figure 2.**
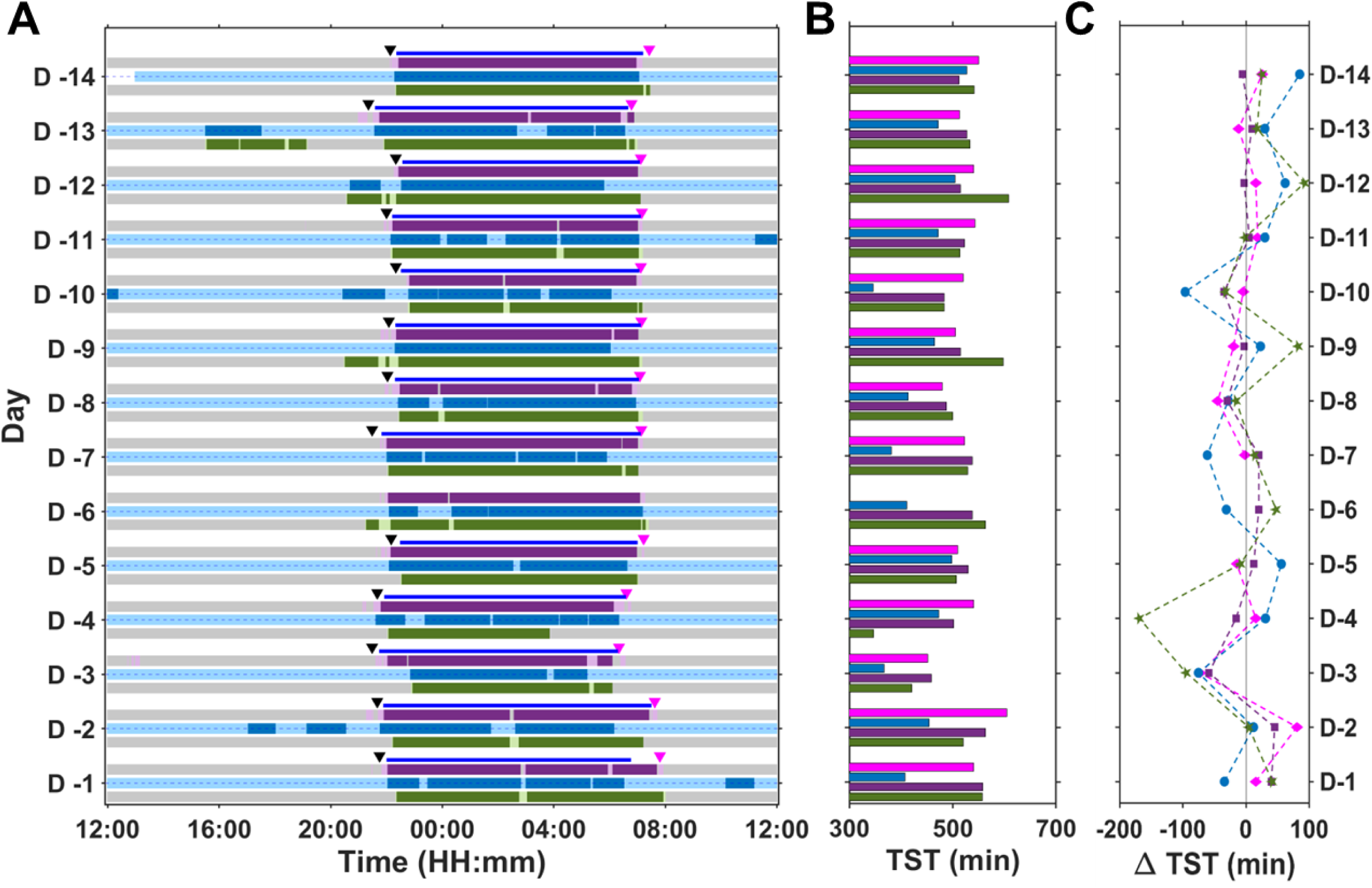
Sleep patterns from a PLWD aged between 70 and 75 collected at home over a period of 14 nights. **A.** The raster on the left depicts the sleep-wake data and the associated Sleep Diary information. The dark coloured regions indicate sleep; light coloured region indicate wake while grey indicates absence of data. The inverted triangles above the coloured regions indicate the get to bed time and get out of bed time from Sleep Diary while the solid navy blue line between the triangles indicates the sleep period time (SPT). **B.** Total sleep time (TST). **C.** Deviations from mean TST. The devices are colour coded; Withings sleep analyser: Purple; Axivity: Blue; Withings watch: Green and Sleep Diary: Magenta.

### Laboratory validation against PSG and at-home between-device associations

Although the primary focus of the current analysis was on the reliability and utility of CST-derived sleep measures, we also examined their validity against the gold standard PSG and the between-device associations of the raw sleep measures. We assessed the sleep summary and sleep stage duration measures derived from Sleep Diary, Axivity, WSA and WW against gold standard polysomnography in the laboratory. There were a total of 65 laboratory nights available for Sleep Diary, 59 for Axivity, 73 for WSA and 65 for WW. The Bland-Altman metrics and scatter plots are provided in Supplemental Table S2 and Figures S1 to S4.

Overall, moderate significant (p<0.001) correlations with PSG were observed for Sleep Diary TST (r =0.38), Axivity TST (r =0.50), WSA TST (r =0.46), WSA WASO (r =0.38) and WSA SEFF (r =0.51). WW had a significant correlation (r=0.27, p=0.028) with PSG only for sleep efficiency. Across all devices, TST and SEFF were overestimated, while WASO was underestimated relative to PSG. WW showed an overall constant SOL of ∼3min while SD and WSA showed more variability in estimation.

None of the sleep stage duration measures obtained by the two Withings devices were significantly correlated with the corresponding PSG measures (Supplemental Figure S5; Table S3). Both devices overestimated deep sleep duration with WSA (Bias: 61.5 min) performing slightly less poorly than WW (Bias: 180.8 min). Light sleep duration was underestimated by WW (Bias: 9.6 min) and overestimated by WSA (Bias: 43.5 min). WW did not provide a REM sleep duration measure while WSA REM sleep duration was overestimated (Bias: 27.1 min). When expressed as a percentage of TST (% TST), the sleep stage duration measures had a similar pattern of concordance (Figure S6). We also examined the association between WSA AHI and PSG AHI and found a strong correlation between them (r =0.76, p<0.001) with a very small bias of -0.4±9.4 events/hour (Figure S7).

Between-device Pearson correlations of sleep measures obtained during the laboratory recording showed a moderate association between Axivity TST and WSA TST (r = 0.52, p <0.001) and a weak association between Sleep Diary SOL and WSA SOL (r = 0.31, p = 0.033). Other summary measures obtained from the various devices and labelled as Sleep Efficiency or WASO did not correlate significantly across devices.

To provide further context for the interpretation of similarly labelled raw sleep measures, we examined between-device associations in the at-home data. Repeated-measures correlations (rmcorr) were computed for the raw sleep measures derived from single nights (Supplemental Table S4). Moderate correlations were observed for several duration and timing measures across the objective sleep monitoring devices. Correlations involving the Sleep Diary and most other between-device associations were weak (r<0.3). Thus, commonly used CST-derived sleep measures such as SOL, SEFF and WASO do not correlate in a meaningful way across devices when correlations are based on single night assessments.

### Reliability of sleep measures

To characterise the reliability of the sleep measures across devices and participant groups, we first quantified reliability using the single-night ICC(1,1) (also referred as single night reliability) and examined how reliability varied by device and sleep aspect. We then evaluated the extent to which reliability improved with multi-night aggregation. ICC varied across devices and groups with most sleep measures having an ICC less than 0.7 (Figure 3A and Supplemental Table S5). Of the different sleep aspects (Supplemental Figure S8A), Duration measures (TST, SPT, Recording period time (RPT), 24-hour Total sleep time (TotalSleep)) had overall higher ICC(1,1) followed by Timing (Get to bed time (BT), Wake time/Get out of bed time (WT), Mid sleep time (MST)) and Continuity measures (WASO, SEFF, NAW, SOL), while nap measures had the lowest ICC (duration of naps (DUR_NAP), number of naps (NNAP))).

**Figure 3.**
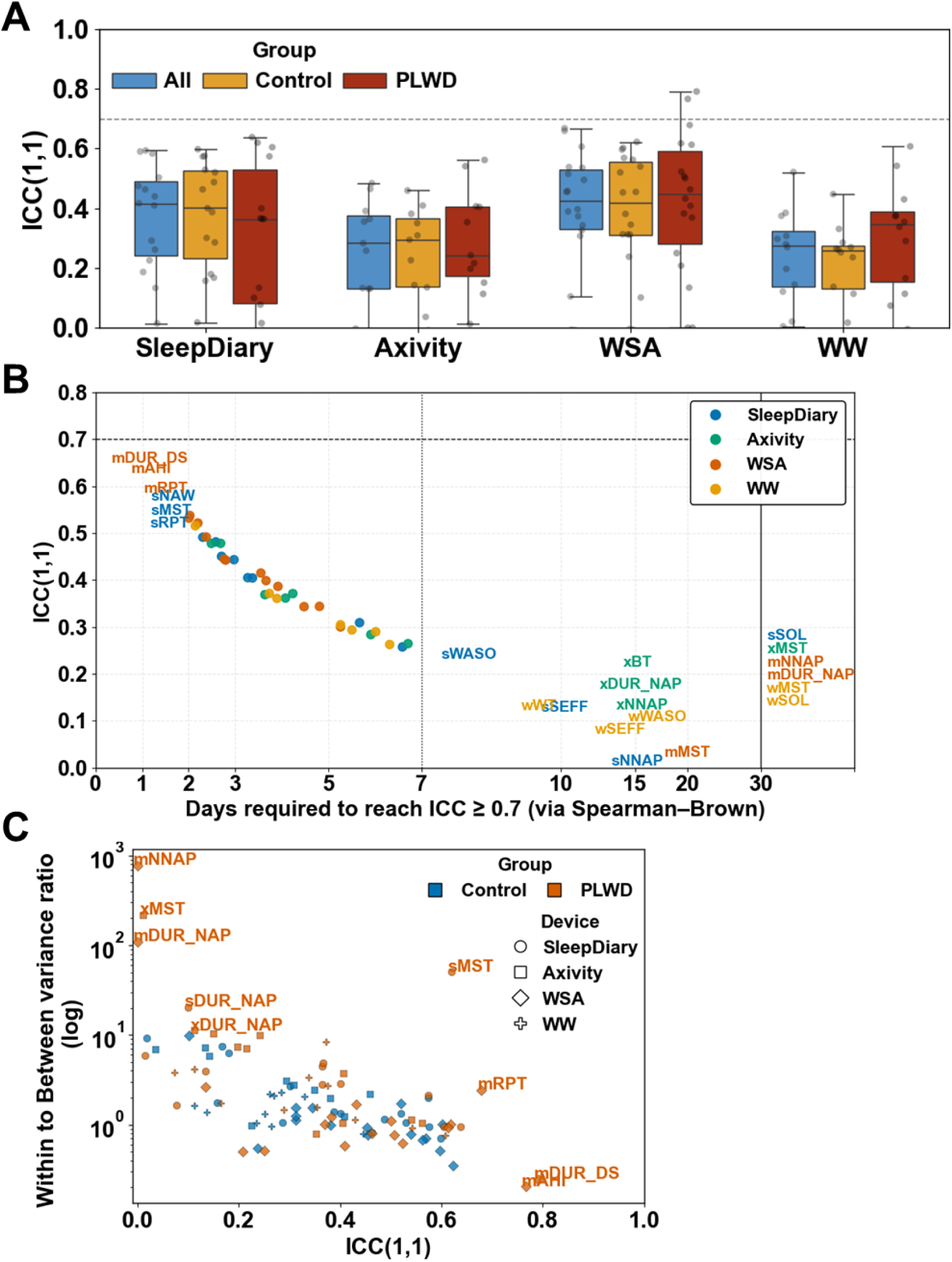
Reliability of sleep measures across devices and participant groups. **A.** Single-night reliability (ICC(1,1)) across devices for all participants, Controls, and people living with dementia (PLWD). **B.** Association between ICC(1,1) and the number of nights of aggregation required to reach acceptable reliability (ICC ≥ 0.7). Measures to the right of the vertical line fail to reach acceptable reliability even after 30 nights. **C.** Association between ICC(1,1) and within-to-between variance ratio of the measures for Controls and PLWD

The number of nights required to achieve acceptable reliability (ICC ≥ 0.7) varied widely across measures with WSA AHI (mAHI), deep sleep duration (mDUR_DS) and time in bed (mRPT) requiring only 2 nights of aggregation (Figure 3B). Seventy-one percent (40/56 measures) of all the sleep measures reached acceptable reliability with seven nights of aggregation (Table 2). Of the sleep measures failing to reach acceptable reliability with seven nights, WW contributed the most (5) followed by Sleep Diary (4), Axivity (4) and WSA (3). With 14 nights (2 weeks) of aggregation, 79% (44/56 measures) reached acceptable reliability (Supplemental Table S6). Measures with low ICC(1,1) consistently required a large number of nights of aggregation to reach acceptable reliability or failed to reach it altogether (Figure 3B).

**Table 2:**
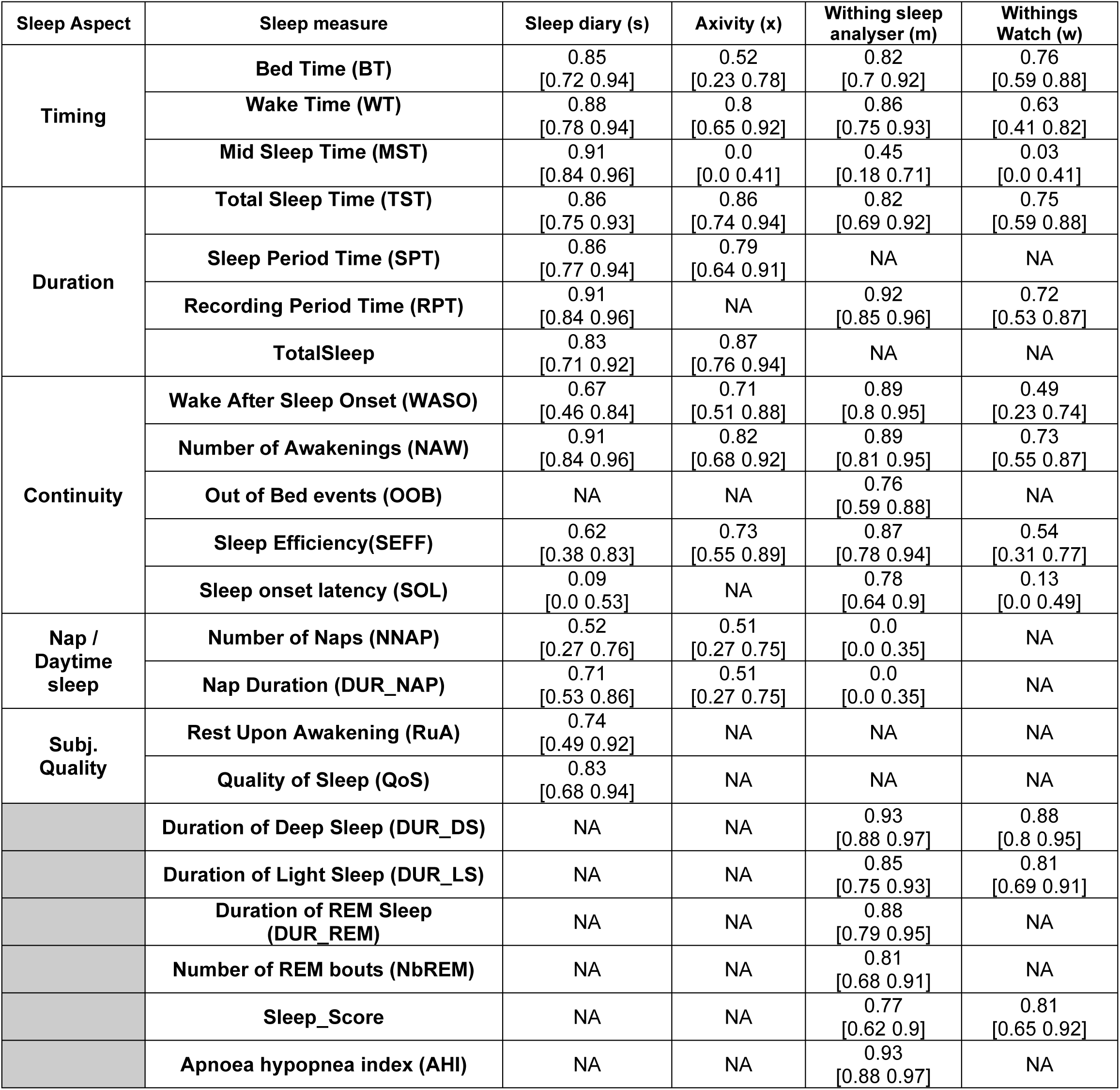
Seven night aggregated reliability for all sleep measures. Values shown are mean followed [95% confidence interval].

To better understand the variance structure underlying these differences and to better characterise why some measures achieved acceptable reliability while others did not, we examined the association between the ratio of within-to-between participant variance and aggregation-related gains in reliability. Figure 3C shows the relationship between ICC and within-to-between participant variance where poor reliability is associated with high within-to-between participant variance ratios. This effect was more marked in PLWD compared to Controls. Across all participants, Controls and PLWD, measures that reached ICC ≥ 0.7 within seven nights exhibited substantially lower within-to-between participant variance ratios compared to those that did not (Figure S8B, Cliff’s delta = -0.81). The cumulative distribution of ΔICC showed that sleep measures failing to reach acceptable reliability exhibited minimal improvement with aggregation, whereas measures that reached ICC ≥ 0.7 demonstrated larger and more consistent improvements (Figure S8C). Further exploration showed that within-to-between participant variability had a strong inverse relationship with ICC (Spearman’s rho = -0.74, p<0.001) and a positive association with ΔICC (Spearman’s rho = 0.69, p<0.001) (Figure S8D). This indicates that aggregation preferentially benefits measures with favourable variance structure (i.e., higher ICC, lower within-to-between ratio, and less within-participant noise).

To determine whether the differences in reliability were primarily driven by the device type or participant group, we modelled the log-transformed within-to-between participant variance ratio using linear mixed-effects models with sleep measures as a random intercept and both device and participant group as fixed effects. In this full model, the reference device (Sleep Diary) had a moderate within-to-between variability (intercept = 0.80, p = 0.014). Relative to Sleep Diary, none of the other devices (Axivity, WSA, or WW) showed statistically significant differences in the within-to-between variability. However, sleep measures from PLWD were associated with a significantly higher within-to-between variance compared to Controls (β = 0.32, p = 0.015), suggesting that participant group contributed more strongly to differences in reliability than device type. When the model was restricted to include only device as a fixed effect, the intercept remained similar but compared with Sleep Diary, WSA exhibited a significantly lower within-to-between variance (β = −0.77, p < 0.001). No significant differences were observed for Axivity or WW relative to Sleep Diary. This suggests that device-related differences in reliability were reduced once participant group was taken into account in the full model.

### Effects of aggregation across 7 nights on between-device associations

Since reliability increases with number of nights recorded, it may be expected that between-device correlations increase when data are averaged over multiple nights. Scatter plots illustrating the effect of single-night versus multi-night aggregation on between-device associations are shown in Figure 4 and the corresponding aggregated correlations are provided in Supplemental Table S4. Aggregation increased the strength of correlations between devices, particularly for Duration and Timing-related measures (e.g., TST, Wake Time, etc.), while improvements were smaller for Continuity measures such as WASO. For sleep stage duration measures such as DUR_DS (single night, r= 0.2; 7 night aggregate, r= 0.25) and DUR_LS (single night, r= 0.3; 7 night aggregate, r= 0.36), the association between WSA and WW did not meaningfully improve with aggregation.

**Figure 4.**
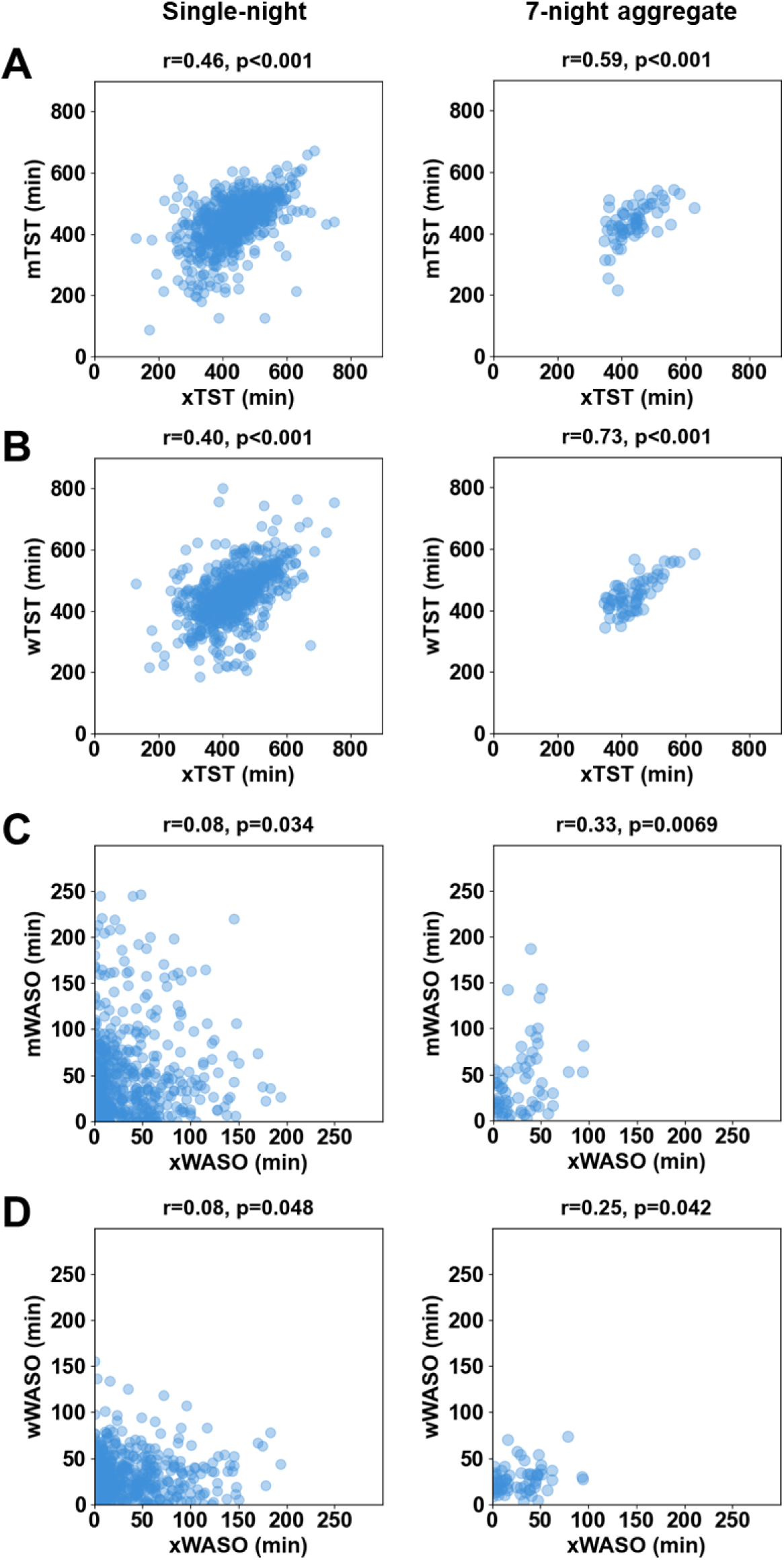
Effect of multi-night aggregation on between-device associations. Scatter plots compare Axivity (x) sleep measures with corresponding measures from Withings sleep analyser (m) and Withings watch (w). A and B. Total sleep time (TST), C and D. Wake after sleep onset (WASO). In each subfigure (A–D), the left panel depicts the association between single-night estimate of the measure, and the right panel shows measure aggregated across 7 nights. Correlation coefficients (r) and p-values are provided at the top of the plots. Correlation type: Single night estimate-Repeated measures Correlation; 7-night aggregates – Pearson’s Correlation.

### Between-device associations for sleep regularity

To examine sleep regularity across devices, Composite Phase Deviation (CPD) was estimated [38]. The reference mean MST was calculated from all available nights for each participant. The single-night phase deviation was then computed relative to the participant-specific mean MST and the previous night’s MST. The corresponding between-device associations are shown in Supplemental Figure S9.

The single-night phase deviation associations were moderate for the objective sleep monitoring devices (Axivity, WSA, and WW; r = 0.28 to 0.32), whereas correlations involving the Sleep Diary were weak (r = 0.08 to 0.12). After aggregation of CPD over 7 nights, all between-device associations improved and were moderate. Correlations between the objective devices and the Sleep Diary ranged from r = 0.38 to 0.42.

### PCA-derived sleep aspects (pSleepAspects)

We next investigated whether the reliability of the sleep aspects within each device and association between devices could be improved by dimensionality reduction of the sleep measures from each device. We first applied PCA followed by Varimax rotation to the raw sleep measures from each device. Across all devices, the raw PCA components were interpretable and accounted for a large proportion of the total variance (75.8 to 84.3%), but the number and composition of components varied by device (Figure S10). PCA-derived sleep aspects (pSleepAspects) such as Duration, Continuity and Timing were identified across all devices. For Sleep Diary, five components (Duration, Continuity, Timing, Naps and Quality) were retained, explaining 75.8% of the variance. For Axivity (Duration, Continuity, Naps and Timing), four components were retained, explaining 84.3% of the variance. For the Withings Sleep Analyzer (WSA), seven components (Continuity, Duration, Naps, Timing, NREM-AHI, REM and Efficiency) were retained, explaining 82.9% of the variance. Here, the NREM-AHI component was positively associated with light sleep duration (mDUR_LS) and AHI and negatively influenced by deep sleep duration (mDUR_DS). For the Withings Watch (WW), four components (Duration, Continuity, Timing and NREM) were retained, explaining 82.3% of the variance.

Following this, PCA was repeated on a reduced set of measures after removing measures that did not reach acceptable reliability (ICC < 0.7 after seven nights of aggregation) (Figure 4A). In this analysis, Duration and Continuity pSleepAspects were identified across objective sleep monitoring devices. In Axivity, the total number of components was reduced to two components explaining 80.8% of the variance. The pSleepAspects were identified as Duration and Continuity. In WSA, five components were retained and identified as Continuity, Duration, NREM-AHI, Efficiency, and REM, explaining 79.5% of the variance. The Withings Watch components were reduced to only two components, Duration and Continuity, explaining 74.1% of the variance. In the subjective tool, Sleep Diary, four pSleepAspects were identified, namely Duration, Quality, Timing, and Naps, explaining 74.7% of the variance. In the Sleep Diary, the continuity measures except for NAW are lost in this analysis due to the subjective measures of WASO, SEFF and SOL failing to reach acceptable levels of reliability with 7 nights of aggregation.

When pSleepAspect were derived from the ‘clean’ set of measures, i.e., after removal of measures that did not reach acceptable reliability, all pSleepAspects reached acceptability within seven nights of aggregation across all devices. In contrast, when pSleepAspects were derived from the full set of raw measures, at least one component per device did not reach the ICC ≥ 0.7 threshold (Figure 4C).

### Associations between PCA-derived Sleep Aspects (pSleepAspects)

To examine how these pSleepAspects related across devices, rmcorr was computed between all pairs of pSleepAspects obtained from the raw and clean PCA analyses and corrected for multiple testing. In both analyses, the Duration-related pSleepAspects showed the most consistent cross-device associations, with moderate positive correlations between Axivity Duration and WSA Duration (Figure 5B and Supplemental Figure S11). Associations were also observed between WW Duration and both WSA and Axivity Duration. Please note that the direction of these correlations reflects the PCA loadings rather than a true inverse relationship. All devices had weak correlations with Sleep Diary Duration. The associations between the Duration-related pSleepAspects were improved with 7-night aggregation(See Supplemental Table S7). The associations based on the PCA were not stronger than those observed for comparable sleep aspects derived from the single-night raw sleep measures (please compare supplemental tables Table S4 and S7).

**Figure 5.**
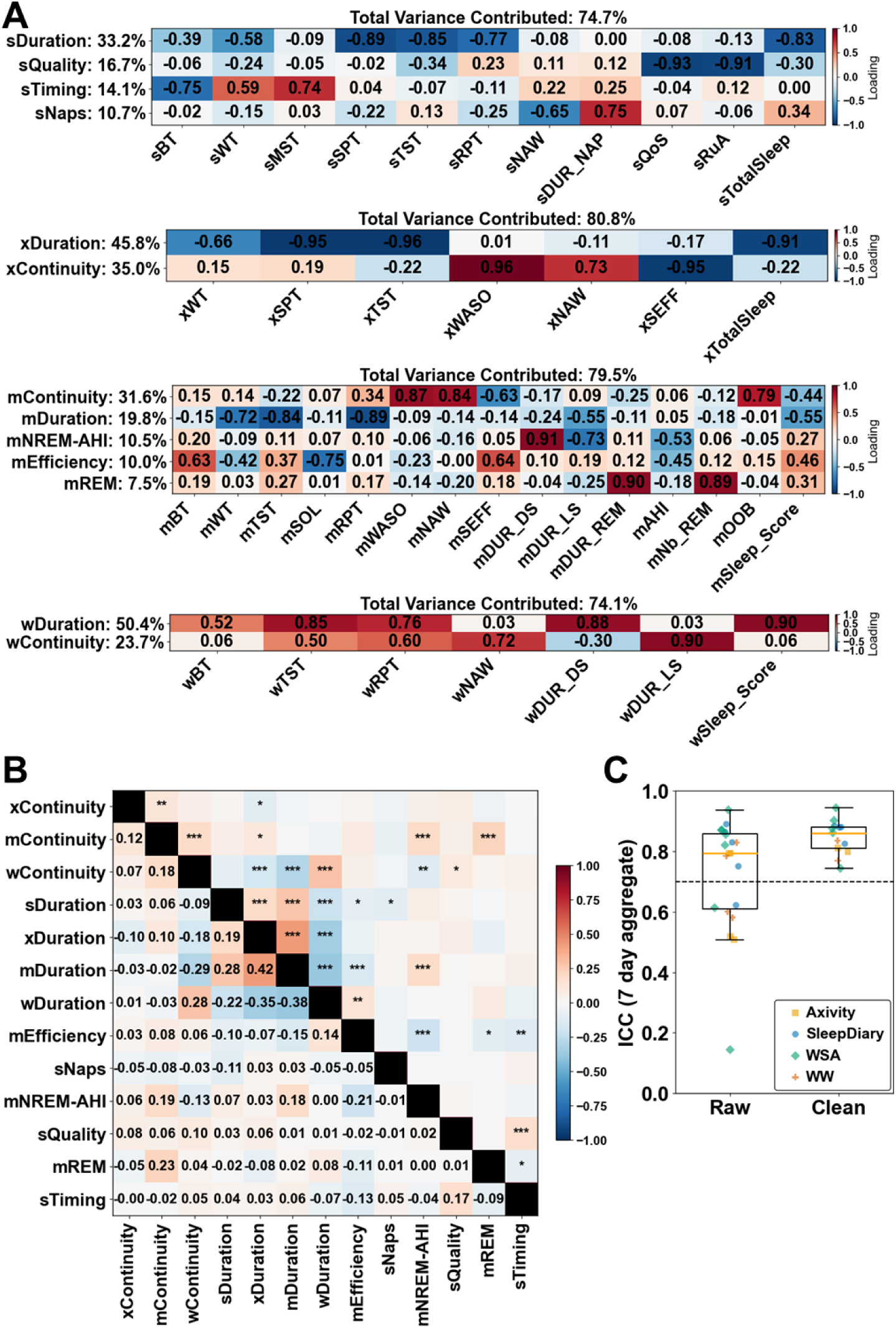
Analysis of sleep aspects across devices. **A.** Principal component analysis (PCA) of sleep measures with the measures that reach acceptable reliability (ICC ≥ 0.7) with 7 nights aggregation. The device components are marked with s – Sleep Diary; x – Axivity; m – Withings sleep analyser (WSA); w – Withings Watch (WW). Identified component labels and the percentage of variance explained are shown on the left of the heatmap of the standardized loadings, and total variance in the device explained components with eigen values over 1 is indicated above each panel. **B.** Heatmap of the association between single-night scores of PCA-derived sleep aspects (PSAs) identified across devices estimated via repeated-measures correlation (left of the diagonal); multiple testing correction was performed using the Benjamini-Hochberg false discovery rate and adjusted q-values (right of the diagonal) are provided; asterisks denote significance levels (* q<0.05, ** q<0.01, *** q<0.001). C. Seven night aggregate of ICC estimated from the PSAs of devices. Raw indicates the PSAs derived using all the sleep measures while Clean indicates the PSAs derived using the measures that fail to reach acceptable reliability (ICC ≥ 0.7) with 7 nights aggregation removed.

In the analyses using all measures, i.e. not just the reliable measures, Timing-related pSleepAspects showed associations between WSA and both Axivity and WW, while Axivity and WW Timing showed a positive correlation. As with Duration, the direction of these correlations arose from the PCA loadings (Supplemental Figure S11). These Timing-related pSleepAspects associations improved the most with aggregation (Supplemental Table S7). This component was not present in the clean analysis since it was associated with unreliable raw timing measures. There were weak correlations between the two Withings devices for Continuity and between WSA Continuity and WW Duration (see Figure 5B).

To further explore the interchangeability of the pSleepAspects between the devices, we pooled the clean sleep measures across all devices and performed the PCA analysis as depicted in Figure 6. The top five PCs were identified as PC1: Subjective and Objective Duration, PC2: Withings Sleep Analyser (WSA), PC3: Sleep Diary, PC4: Subjective and Objective Timing and PC5: Objective Axivity Continuity. These components follow the results of the association analysis with Duration and Timing being the only components shared across all devices (both subjective and objective). The remaining of the components were primarily driven by individual devices. The WSA component had weak associations with the WW component, as also observed in the cross-device association analysis.

**Figure 6.**
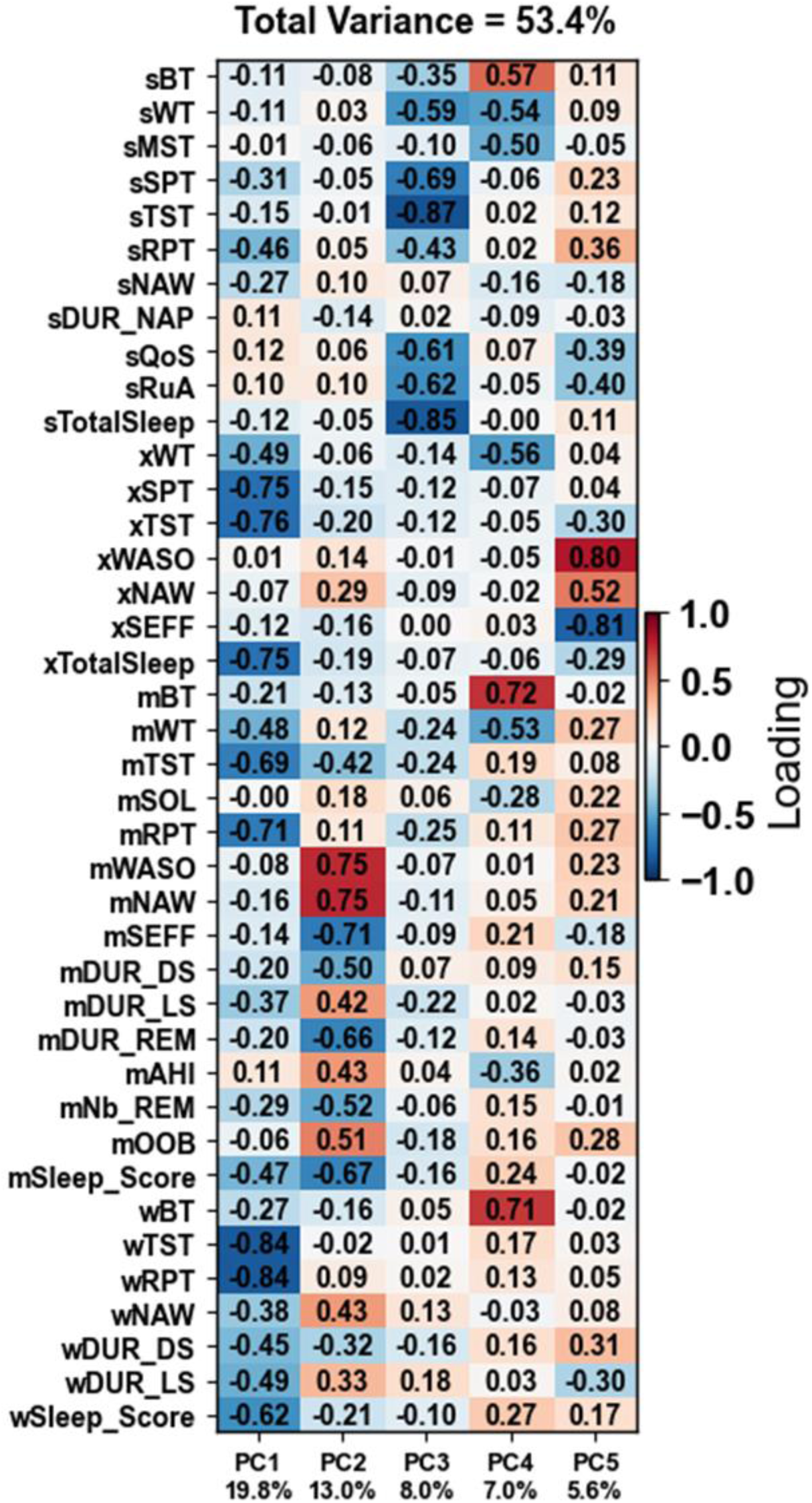
Principal component analysis (PCA) of sleep measures pooled across all the four devices. The measures that fail to reach acceptable reliability (ICC ≥ 0.7) with 7 nights aggregation removed. The top five PCs were identified as PC1: Subjective and Objective Duration; PC2: Withing Sleep Analyser; PC3: Sleep Diary; PC4: Subjective and Objective Timing; PC5: Objective Axivity Continuity.

### Predictive performance of the sleep measures

We next investigated whether CST measures and pSleepAspects could be used to discriminate between PLWD and Controls. We first fitted models using the raw sleep measures from each individual devices and all raw sleep measures pooled across devices (Supplemental Table S8). We then evaluated models based on the raw and clean pSleepAspects. The discriminative performance of the devices varied across devices, with different sleep measures contributing to the performance of the devices. Among individual devices, the Sleep Diary and Axivity based models showed moderate discrimination between PLWD and Control with an AUC of 0.64 and 0.59, respectively.

This was followed by the Withings Watch (AUC = 0.50) and was weakest for the Withings Sleep Analyzer (AUC = 0.44). The highest overall performance was achieved in the pooled dataset, i.e. when all available data from all devices were used (AUC = 0.70). Across all the models, AP values were generally lower than AUC, followed a similar trend to AUC, and were more variable across cross-validation folds compared with AUC. SHAP analyses of the models revealed the measures that drive the performance and showed that the relative importance of variables varied across devices (Supplemental Figure S12). In the pooled analysis, the top five sleep measures that contributed to the discrimination between Controls and PLWD were Sleep Diary and Axivity derived duration and continuity measures.

We next analysed the discriminative power using PCA derived sleep aspects either based on all variables (raw) or only using reliable (clean) variables. When models were trained using PCA derived Sleep Aspects, Axivity showed the clearest improvements in both AUC and AP for clean compared with raw pSleepAspects, while WSA showed smaller improvements (Table 3). Overall, performance of both Axivity and WSA was higher with the clean pSleepAspects compared to using raw pSleepAspects. WW did not show any improvements with the pSleepAspects. The Axivity model with clean pSleepAspects achieved higher performance (AUC = 0.71) than the pooled raw measures model. For Sleep Diary, removing low-reliability components led to a slight reduction in both AUC and AP but led to more consistent performance across cross-validation folds.

**Table 3.**
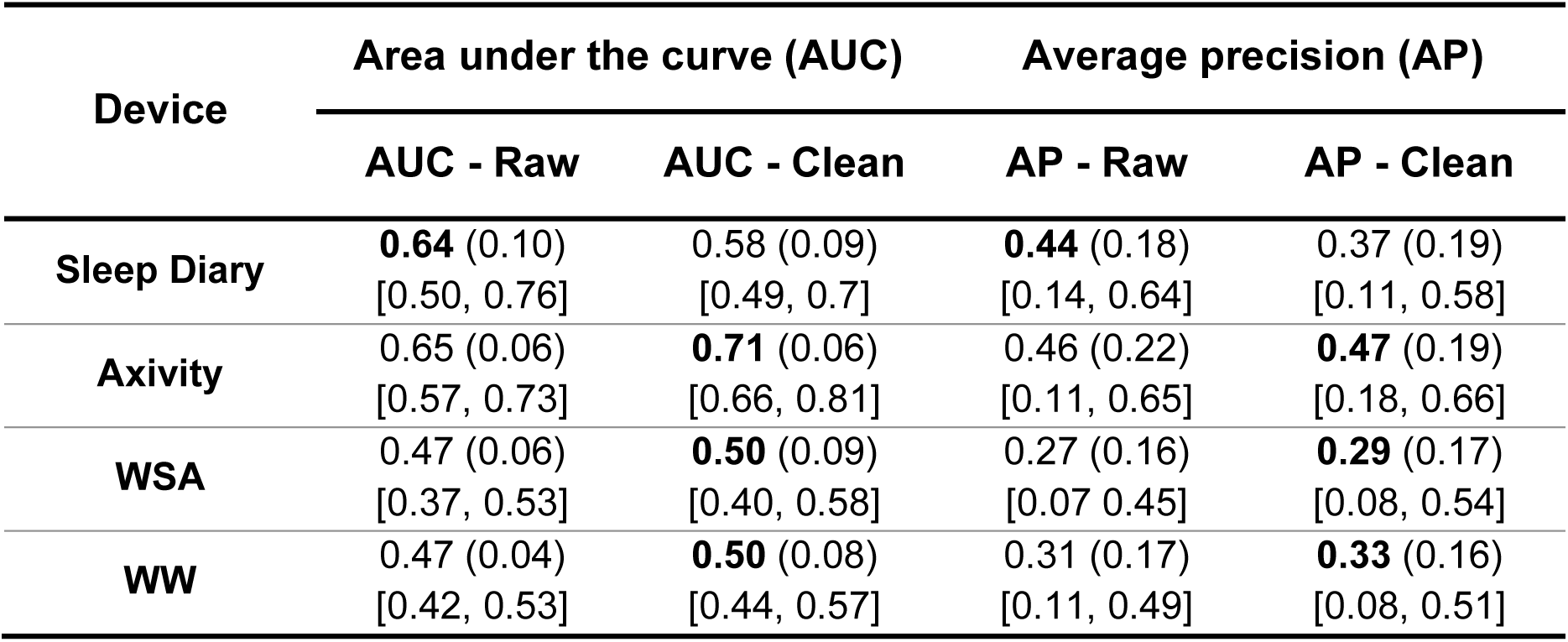
Discriminative performance of the devices – PCA derived sleep aspects. Note: The values shown are the mean followed by the (standard deviation) of 5-fold cross validation and [min max]. Raw indicates the PCA derived sleep aspects (pSleepAspects) obtained using all the sleep measures while Clean indicates the pSleepAspects derived using the measures that fail to reach acceptable reliability (ICC ≥ 0.7) with 7 nights aggregation removed.

Finally, we compared the overall contribution (mean absolute SHAP values) of the sleep measures to the discriminative performance with their single-night reliability (ICC(1,1)) for all the devices and the associations are depicted in Supplemental Figure S13. The correlation between the single-night reliability and the mean absolute SHAP values was strongest and statistically significant for WSA (Spearman’s rho = 0.63, p=0.005), moderate for Sleep Diary (Spearman’s rho = 0.44), WW (Spearman’s rho = 0.45), and Axivity (Spearman’s rho = 0.39).

## Discussion

In this work, we assess the comparability and utility of sleep measures from two consumer sleep technologies, research grade actigraphy, and a sleep diary. We first show the poor agreement between CST and PSG measures of sleep and that only a few identically labelled raw sleep measures were associated between devices. We next applied a methodology to identify and filter CST-derived sleep measures based on night-to-night stability to assess their utility in detecting non-PSG aspects of sleep in longitudinal use. The methodology was applied to simultaneously collected at-home data from four devices/modalities in a group of older adults and PLWD. Bivariate correlations between variables with similar names were in general weak, with stronger associations mainly limited to some Duration- and Timing-related measures between objective devices. To investigate whether measures at least could detect individual specific aspects of sleep, we conducted a reliability analysis. The majority of automatically generated sleep measures reached acceptable reliability with seven nights of aggregation, but a substantial subset remained unstable even after two weeks of aggregation and the number of nights required was larger in PLWD than Controls. These findings highlight marked variation in measures and their night-to-night stability across CSTs despite their similar and PSG-derived terminology. PCA applied separately to each of the devices using measures that demonstrated acceptable reliability, yielded reliable latent sleep aspects, with components that can be interpreted as Duration and Continuity across all CSTs. The Duration- and Timing-related PCA-derived sleep aspects were the only component that had moderate between-device association implying that even after dimensionality reduction different devices in general measure different aspects of sleep. Both in the raw sleep measures and the PCA-derived sleep aspects Timing-related measures showed the most improvement in reliability and between-device associations. Finally, investigating to what extent sleep related measures from various devices may be useful in creating a digital biomarker to distinguish between Controls and PLWD we demonstrate that filtering out unstable measures and using PCA derived sleep aspects from the remaining stable measures (ICC≥0.7) can improve predictive performance and/or reduce fold to fold variability in model performance for Axivity and WSA. However, the relative contribution of sleep aspects to discrimination between PLWD and Controls varied across devices. Overall, these results indicate that, with the exception of sleep duration measures, different devices often measure different aspects of sleep even though the variables have identical or near identical labels. Nevertheless, CSTs may be useful for characterising sleep and for the identification of biomarkers, but only when their use is guided by measure-specific reliability, adequate multi-night aggregation, and a careful avoidance of assuming interchangeability across devices.

### Laboratory Validation

Validation against laboratory PSG reinforced well-known limitations and clarified what the CST-derived sleep measures do and do not represent. Weak significant associations with PSG were observed only for Sleep Diary TST, Axivity TST, and WSA TST, WASO, and SEFF, whereas WW showed no significant correlation with PSG summary measures. In addition, both Withings devices showed poor concordance for sleep stage duration measures, with overestimation of deep sleep (DUR_DS vs N3) and no meaningful association with PSG-derived REM sleep and Light sleep (N1+N2) durations. These findings are consistent with the broader literature showing that non-EEG CSTs can only estimate sleep duration and timing related aspects of PSG sleep and do not provide a meaningful estimate of NREM and REM sleep stages [39, 40].

### Reliability and effects of aggregation

Despite the limited concordance with PSG, these CST-derived sleep measures may nevertheless be useful for characterising sleep-related behavioural/physiological states, provided they are sufficiently reliable across nights. Reliability is important because a useful sleep measure should ideally reflect stable differences in an individual’s latent sleep that are captured by the CST rather than within-participant night-to-night noise or algorithmic instability. In our analysis, the majority of sleep measures reached acceptable reliability with multi-night aggregation, with 71% reaching ICC ≥ 0.7 within seven nights and 79% within 14 nights or 2 weeks of aggregation. However, across devices and sleep aspects, a substantial subset of the raw sleep measures remained unstable even after a week of aggregation, and even after 2 weeks some measures still did not reach acceptable reliability. This finding implies that the number of nights needed depends strongly on the specific measure used and the characteristics of the population that is investigated. In our data, many duration-related measures approached a reliable trait-like estimate within a week of aggregation, while continuity and nap-related measures often required a longer aggregation window or failed to stabilise even with two weeks of aggregation. This is directly relevant to study designs where CSTs are used to estimate sleep measures. For example, if the aim is to get an estimate of a participant’s habitual sleep duration for longitudinal monitoring, relatively few nights would be sufficient. In contrast, if the goal is to quantify sleep fragmentation such as number of awakenings or sleep efficiency, substantially longer monitoring period may be required. This is consistent with actigraphy guidance recommending multi-night recordings, often spanning several days to weeks depending on the application, and with large-scale cohort studies such as UK Biobank, which used 7-day Axivity recordings to derive habitual behavioural measures [39, 40]. The current analyses also shows that reliability may be more difficult to achieve in populations such as PLWD and older adults, although the number of nights required still depended strongly on the specific measure [41].

The variance-structure analyses helped explain why some measures stabilised with aggregation whereas others did not. Measures with poor ICC had high within-to-between participant variance ratios, and those failing to reach acceptable reliability showed minimal gains with aggregation. This implies that simply recording more nights will not necessarily improve all CST-derived measures. Aggregation is beneficial only when night-to-night variation is dominated by random within-person physiological noise that reduces with averaging, but it is less useful when a measure contains substantial device- or algorithm-specific instability. This is important for longitudinal study design, because it helps to identify measures that are suitable for estimating habitual sleep characteristics and those that are unlikely to provide stable individual-level estimates even with longer monitoring. Once participant group was included in our mixed-effects model analysis, device effects were no longer statistically significant, whereas PLWD showed significantly higher within-to-between participant variance than Controls. This suggests that differences in reliability are not just about which device is being used, but also reflect the population being monitored. In the context of older adults and PLWD, where sleep may be inherently more fragmented, higher within-person variability may be expected and may itself limit the usefulness of some CST-derived measures as longitudinal outcomes due to their inherent lack of specificity in measuring quiet wake.

### Latent Sleep Aspects

When PCA was used to derive PCA-derived sleep aspects (pSleepAspects), i.e., latent CST components representing broader sleep domains such as Duration, Continuity and Timing, the resulting structure was sensitive to the inclusion of unstable measures [2, 36]. Within the measurement framework for estimating the latent sleep state, CST-derived sleep measures are considered as device-specific, imperfect representations of the latent sleep. Here, PCA serves to identify a broader latent structure within each device’s representation of sleep, but the extent to which these components are meaningful depends on the reliability of the input measures. In the PCA derived from the full set of raw sleep measures, the components were broadly interpretable and captured substantial variance, yet at least one component per modality failed to reach acceptable multi-night stability, indicating that some of the recovered structure was partly driven by unstable variance rather than trait-like sleep characteristics. Restricting PCA to measures that achieved acceptable reliability produced a more stable latent representation, with the resulting “clean” pSleepAspects consistently reaching acceptable ICC across devices. This supports the role of reliability filtering as a quality control step towards extracting reliable multidimensional sleep constructs and improves confidence that the resulting components reflect stable aspects of sleep rather than measurement error. However, the concordance analyses revealed that the PCA approach only led to marginal improvements in across device concordance of sleep quantification.

The device or tool-specific outcomes of the reliability filtering are also informative for selecting which measures from a specific tool are suitable for longitudinal monitoring and which are not. In Sleep Diary, several continuity-related subjective measures did not survive reliability filtering, with the PCA decomposition dominated by duration, timing, naps, and subjective sleep quality. Practically, this means that short diary protocols may be more defensible for estimating habitual self-reported sleep duration or bed/wake timing than for estimating nocturnal awakenings or other continuity measures [42].

### Duration measures

Across our analyses, “Duration” emerged as the only sleep aspect that is stable, consistent and interchangeable between the objective sleep monitoring devices and can act as a robust digital sleep biomarker. This was evident in the repeated-measures correlation analysis of pSleepAspects, where Duration pSleepAspects showed consistent but moderate between device associations, whereas Continuity pSleepAspects were weakly associated. This means that although there are observed differences in the absolute estimate of the various duration measures (e.g., total sleep time, time in bed etc.,), the interchangeability is better supported at the level of identifying relative changes and stable within-participant differences. This finding is notable since CST derived total sleep time and related duration measures have previously been reported as outcome measures in several longitudinal digital health studies [43, 44].

### Device interchangeability

The pooled PCA on the clean sleep measures reinforced this conclusion with Duration (and Timing) appearing as the top component shared across all devices followed by device-specific components having an equally large portion of variance. Considered together, these findings suggests that Duration-related measures, may be useful as candidate biomarkers because they are comparatively stable, interpretable, and modestly associated between devices. However, the pooled PCA also showed that other pSleepAspects remained device-specific indicating the need for caution against treating nominal sleep measures derived by different sensing modalities and physiological proxies as equivalent across devices or to PSG. This finding is important since CST derived sleep stage duration and continuity measures are increasingly used as “sleep” variables in large observational studies with an implied connotation of these measures being equivalent to PSG sleep duration despite known lack of population-specific validation and lacking clear demarcation that the measures are device-specific [45, 46].

### Discriminative Utility

We also used PLWD-versus-control discrimination as an example to illustrate how these measurement properties affect downstream predictive model development. The models based on the raw sleep measures served as a baseline for the follow-up models using the raw and clean pSleepAspects. Clean (reliability filtered) pSleepAspects improved performance and/or reduced fold-to-fold variation for Axivity and WSA with Axivity showing the clearest improvement. Variability in fold-to-fold performance is caused partly by instability/noise in the underlying measures, where measures with low reliability (high within-to-between participant variance) can yield spurious patterns that improve discrimination in one split but fail to generalise to another. It is noteworthy that the lower and more variable AP compared to AUC is due to AP being sensitive to specificity of the positive prediction (PLWD) across folds and penalises the misclassification of PLWD more stringently. Although our data set is limited, this reinforces the importance of model stability and need for reliability (ICC or within-to-between participant ratio) feature selection and reporting in CST based predictive modelling. We believe that incorporating a reliability driven feature selection and reporting as a routine step will to improve model stability and between-cohort model generalisation which are well-known gaps in CST based digital health models across populations [47]. Finally, we observed that there was a general positive correlation between feature contribution (mean absolute SHAP values) and reliability (ICC(1,1)) across all devices, which further supports a link between stability and predictive value. This should be interpreted as evidence that reliability of sleep measures improves model behaviour, rather than as a claim on validity of the reliable features as physiologically accurate and equivalent to PSG measures.

## Conclusion

CSTs are increasingly deployed for longitudinal sleep monitoring in older adults and PLWD and the sleep measures generated by them are frequently treated as equivalent to PSG measures despite their proprietary algorithms, limited transparency, and varying validity across populations. In this work, we show that night-to-night stability is a key prerequisite for the utility of CST-derived sleep measures in longitudinal research where comprehensive PSG validation is impractical. Although many sleep measures reached acceptable reliability with multi-night aggregation, a substantial (∼30%) subset remained unstable even after one to two weeks of averaging. Aggregation also improved between-device associations for duration and timing related raw measures, whereas improvements for continuity-related measures were limited. Filtering measures based on acceptable reliability before dimensionality reduction yielded stable and interpretable sleep aspects, i.e., latent sleep domains such as Duration, and Continuity. Between-device association analysis revealed that Duration was the only sleep aspect that was consistently shared across objective sleep monitoring devices, with most other measures remaining device-specific and non-interchangeable. Overall, these findings support a measurement reliability-aware approach to digital sleep phenotyping in older adults and PLWD. CST-derived measures should not be assumed to be interchangeable across devices or equivalent to PSG based on shared labels alone. Instead, the practical utility of CST sleep measures as sleep biomarkers depends on three factors: how well they agree with gold standard PSG, how many nights are needed before they behave like a stable trait-like measure, and whether it captures an aspect of sleep that is interchangeable across devices. Overall, these findings support for designing longitudinal sleep studies around the required reliability of the target outcome, rather than on a single default monitoring period.

## Supporting information

The supplementary materials comprises of supplementary tables, and figures that support and extend the findings presented in the main text.

## Acknowledgement

This work was primarily supported by the UK Dementia Research Institute, Care Research & Technology Centre at Imperial College, London and the University of Surrey, Guildford, United Kingdom [award number: UKDRI-7206 and PP202216] which receives its funding from UK DRI Ltd, funded by the UK Medical Research Council, Alzheimer’s Society and Alzheimer’s Research UK. In part supported by NIHR Oxford Health Biomedical Research Centre [NIHR 203316]. The authors thank the staff members Damion Lambert, Tegan Ward, Jasmine Ramble and the members of the Clinical Research Facility for their help with data collection and curation. They would also like to extend their thanks to the members of Surrey and Borders Partnership (SABP) for help with the recruitment and screening of participants.

KR and DJD envisioned the research question. KR conceived the methodology, conducted the data exploration and analysis and prepared the manuscript. DJD, CdM, GA, HH, RN and VR contributed to the design of the studies, and were responsible for participant recruitment and screening, and study conduct. HH served as the Clinical Lead/ Chief Investigator in Study 2 (NHS ethics: 22/LO/0694), taking responsibility of ethics submission, participant’s safety, and all clinical procedures. GA, MMP and CdM scored the PSG records and created the consensus hypnogram. All the authors contributed to finalising the manuscript.

## Data Availability

The data used in this study are available from the co-author Ciro della Monica upon reasonable request. Contact.

