## Supplementary material for "Are different consumer sleep technologies measuring the same essential aspects of sleep?": The supplementary materials comprises of supplementary tables, and figures that support and extend the findings presented in the main text.

Name: Dr Kiran Kumar Guruswamy Ravindran

Address: Surrey Sleep Research Centre, University of Surrey, GU2 7XP

**Table S1: List of Sleep measures**

The Withings API reference (https://developer.withings.com/api-reference/) contains the exact definitions of the Withings measures described here. Presence (Y/N) of the measures and device specific note is given within brackets '()' in the device columns. The timing measures are expressed as fractional hours over a 24-hour period.

| Sleep Aspect | Sleep measure | Unit | Definition | Sleep diary (s) | Axivity (x) | Withing sleep analyser (m) | Withings Watch (w) |
|---|---|---|---|---|---|---|---|
| Timing | **Bed Time (BT)** | Fractional hours | Time get to bed or start of recording period | Y | Y | Y | Y |
| | **Wake Time (WT)** | Fractional hours | Time get out of bed or end of recording period | Y | Y | Y | Y |
| | **Mid Sleep Time (MST)** | Fractional hours | Mid-point between BT and WT | Y | Y | Y | Y |
| Duration | **Total Sleep Time (TST)** | minutes | Total time spent asleep | Y | Y | Y | Y |
| | **Sleep Period Time (SPT)** | minutes | Time spent from sleep onset to final awakening | Y | Y | N | N |
| | **Recording Period Time (RPT)** | minutes | Total time in bed or recording period determined by device | Y | N | Y | Y |
| | **TotalSleep** | minutes | Sum of all sleep bouts in a 24 hour period | Y | Y | N | N |
| Continuity | **Wake After Sleep Onset (WASO)** | minutes | Time spent awake after falling asleep | Y | Y | Y | Y |
| | **Number of Awakenings (NAW)** | count | Number of times the participant wakes up | Y | Y | Y | Y |
| | **Out of Bed events (OOB)** | count | Number of times the participant left the bed | N | N | Y | N |
| | **Sleep Efficiency(SEFF)** | % | Ratio of TST to the RPT/xSPT expressed as a percentage | Y | Y | Y | Y |
| | **Sleep onset latency (SOL)** | minutes | Time taken to fall asleep | Y | N | Y | Y |
| Nap / Daytime sleep | **Number of Naps (NNAP)** | count | Total number of naps | Y | Y | Y | N |
| | **Nap Duration (DUR_NAP)** | minutes | Sum of the duration of naps | Y | Y | Y | N |
| Subj. Quality | **Rest Upon Awakening (RuA)** | NA | Feeling of restful sleep | Y | N | N | N |
| | **Quality of Sleep (QoS)** | NA | Subjective quality of sleep | Y | N | N | N |
| | **Duration of Deep Sleep (DUR_DS)** | minutes | Duration of Deep sleep (device reported equivalent to N3) | N | N | Y | Y |
| | **Duration of Light Sleep (DUR_LS)** | minutes | Duration of Light sleep (device reported equivalent to N1/N2) | N | N | Y | Y |
| | **Duration of REM Sleep (DUR_REM)** | minutes | Duration of rapid eye movement (REM) sleep | N | N | Y | N |
| | **Number of REM bouts (NbREM)** | count | Number of REM bouts | N | N | Y | N |
| | **Sleep_Score** | NA | Proprietary measures provided by the device | N | N | Y | Y |
| | **Apnea hypopnea index (AHI)** | Events/hour | Average number of hypopnea and apnea events per hour | N | N | Y | N |

**Table S2: Agreement metrics for Sleep summary measures from Sleep Diary (SD), Axivity (AX), Withings sleep analyser (WSA) and Withings Watch (WW) compared to Polysomnography (PSG) in Lab**

Number of participants, N=65 for SD, N=59 for AX, N = 73 for WSA and N=65 for WW. WW does not provide REM sleep duration. The values shown are the mean followed by the (standard deviation) and [95% confidence interval]. The metrics include bias-difference in measurement between the device and PSG; lower and upper bounds of the bias; minimum detectable change (MDC)—smallest detectable change independent of measurement error (half of Bland–Altman agreement width); Standardized absolute difference (SAD)-directionless version of Cohen's d; symmetric mean absolute percentage error (SMAPE)-mean error in measurement expressed as percentage.

| Sleep Measures | | Device Mean (SD) | PSG Mean (SD) | Bias [95% CI] | LoA Lower bound [95% CI] | LoA Upper bound [95% CI] | MDC | SAD [95% CI] | SMAPE [95% CI] |
|---|---|---|---|---|---|---|---|---|---|
| Total Sleep Time (TST) [minutes] | SD | 412.5 (94) | 354.1 (75.1) | 58.5 (95.6) [34.8 82.2] | -128.9 [-169.6 -88.2] | 245.9 [205.2 286.6] | 187.4 | 1.1 [1.1 1.3] | 12 [12 14.4] |
| | AX | 425.7 (121.5) | 360.3 (76.6) | 65.3 (106.7) [37.5 93.2] | -143.9 [-191.7 -96.1] | 274.6 [226.7 322.4] | 209.2 | 1 [1 1.3] | 14.3 [14.3 17.3] |
| | WSA | 487.5 (67.8) | 355.4 (74.5) | 132.1 (74.1) [114.8 149.4] | -13.1 [-42.8 16.6] | 277.3 [247.6 307.0] | 145.2 | 1.9 [1.9 2.2] | 16.9 [16.9 18.9] |
| | WW | 481.2 (93.4) | 364.0 (67.0) | 117.3 (118.2) [88.0 146.6] | -114.5 [-164.8 -64.2] | 349.0 [298.7 399.3] | 231.7 | 1.8 [1.8 2.1] | 17.8 [17.8 20.2] |
| Wake After Sleep Onset (WASO) [minutes] | SD | 38.2 (41.8) | 140.7 (53) | -102.5 (58.1) [-116.9 -88.1] | -216.4 [-241.1 -191.7] | 11.5 [-13.3 36.2] | 113.9 | 2.2 [2.2 2.4] | 62.3 [62.3 69.4] |
| | AX | 23.5 (29.7) | 139.6 (53.8) | -116.1 (57.9) [-131.2 -101] | -229.6 [-255.5 -203.7] | -2.5 [-28.5 23.4] | 113.5 | 2.7 [2.7 3] | 74.8 [74.8 81.9] |
| | WSA | 73.6 (50.3) | 139.0 (53.7) | -65.4 (57.9) [-78.9 -51.9] | -178.9 [-202 -155.7] | 48.1 [24.9 71.2] | 113.5 | 1.4 [1.4 1.7] | 37.2 [37.2 42.4] |
| | WW | 40.5 (27) | 134.4 (51.2) | -93.9 (58.6) [-108.4 -79.4] | -208.8 [-233.7 -183.8] | 21.0 [-3.9 46.0] | 114.9 | 2.4 [2.4 2.7] | 56.8 [56.8 62.6] |
| Sleep Onset Latency (SOL) [minutes] | SD | 25.3 (28.1) | 16 (18.9) | 9.3 (34) [0.9 17.7] | -57.3 [-71.8 -42.9] | 75.9 [61.4 90.3] | 66.6 | 0.8 [0.8 1.1] | 42.9 [42.9 50.3] |
| | WSA | 28.2 (19.4) | 14.5 (17.7) | 13.7 (23.5) [8.2 19.2] | -32.3 [-41.7 -22.9] | 59.7 [50.3 69] | 46.0 | 1.1 [1.1 1.3] | 48.9 [48.9 55.6] |
| | WW | 3.0 (3.6) | 13.0 (11.9) | -10.0 (12.5) [-13.1 -6.9] | -34.4 [-39.7 -29.1] | 14.4 [9.1 19.7] | 24.4 | 1.3 [1.3 1.5] | 58.2 [58.2 65.3] |
| Sleep Efficiency (SEFF) [%] | SD | 86.3 (14.2) | 68.8 (12.6) | 17.5 (17.4) [13.2 21.8] | -16.6 [-24 -9.2] | 51.5 [44.1 58.9] | 34 | 1.6 [1.6 1.8] | 14.3 [14.3 17.1] |
| | AX | 94.8 (6.6) | 69.3 (12.7) | 25.6 (12.9) [22.2 28.9] | 0.3 [-5.5 6.1] | 50.8 [45 56.6] | 25.3 | 2.5 [2.5 2.8] | 16.2 [16.2 18.7] |
| | WSA | 84.5 (8.9) | 69.4 (12.7) | 15.1 (11.2) [12.5 17.7] | -6.8 [-11.3 -2.4] | 37.1 [32.6 41.5] | 21.9 | 1.5 [1.5 1.7] | 11.3 [11.3 13.0] |
| | WW | 91.9 (4.5) | 70.8 (11.2) | 21.2 (11.6) [18.3 24.0] | -1.7 [-6.6 3.3] | 44.0 [39.0 48.9] | 22.8 | 2.5 [2.5 2.8] | 13.6 [13.6 15.6] |

**Table S3: Agreement metrics for sleep stage summary measures from Withings sleep analyser (WSA) and Withings Watch (WW) compared to Polysomnography (PSG) in Lab**

Number of participants, N = 73 for WSA and N=65 for WW. WW does not provide REM sleep duration. The values shown are the mean followed by the (standard deviation) and [95% confidence interval]. The metrics include bias-difference in measurement between the device and PSG; lower and upper bounds of the bias; minimum detectable change (MDC)—smallest detectable change independent of measurement error (half of Bland–Altman agreement width); Standardized absolute difference (SAD)-directionless version of Cohen's d; symmetric mean absolute percentage error (SMAPE)-mean error in measurement expressed as percentage

| Sleep Measures | | Device Mean (SD) | PSG Mean (SD) | Bias [95% CI] | LoA Lower bound [95% CI] | LoA Upper bound [95% CI] | MDC | SAD [95% CI] | SMAPE [95% CI] |
|---|---|---|---|---|---|---|---|---|---|
| Deep (N3) Sleep Duration (DUR_DS) [minutes] | WSA | 145 (67.1) | 83.5 (38.9) | 61.5 (70.5) [45.0 78.0] | -76.8 [-105 -48.5] | 199.8 [171.5 228] | 138.3 | 1.4 [1.4 1.7] | 35.6 [35.6 40.8] |
| | WW | 266.8 (73) | 86.0 (38.9) | 180.8 (76.3) [161.9 199.7] | 31.2 [-1.3 63.7] | 330.4 [297.9 362.9] | 149.6 | 3.1 [3.1 3.4] | 51.4 [51.4 55.8] |
| Light (N1+ N2) Sleep Duration (DUR_LS) [minutes] | WSA | 263.2 (76.5) | 219.7 (60.4) | 43.5 (101.6) [19.8 67.2] | -155.6 [-196.3 -114.9] | 242.5 [201.9 283.2] | 199.1 | 1.3 [1.3 1.6] | 18.6 [18.6 21.4] |
| | WW | 214.4 (85.8) | 224.1 (57.0) | -9.6 (101.9) [-34.9 15.6] | -209.4 [-252.7 -166.0] | 190.1 [146.7 233.5] | 199.7 | 1.1 [1.1 1.3] | 19.1 [19.1 23.4] |
| REM Sleep Duration (DUR_REM) [minutes] | WSA | 79.2 (57.1) | 52.1 (25.2) | 27.1 (57.7) [13.7 40.6] | -86.0 [-109.1 -62.9] | 140.3 [117.2 163.4] | 113.1 | 1.1 [1.1 1.3] | 37.0 [37.0 43.0] |
| DUR_DS (% TST) | WSA | 29.6 (12.9) | 23.7 (10.3) | 5.8 (15.9) [2.1 9.6] | -25.3 [-31.7 -19] | 37 [30.6 43.4] | 31.2 | 1.2 [1.2 1.5] | 29.4 [29.4 34.3] |
| | WW | 56.4 (14) | 23.7 (10.2) | 32.7 (16.1) [28.7 36.7] | 1.1 [-5.7 8] | 64.3 [57.5 71.2] | 31.6 | 2.7 [2.7 2.9] | 41.6 [41.6 46.5] |
| DUR_LS (% TST) | WSA | 54.7 (15.9) | 61.9 (11.4) | -7.2 (18.1) [-11.5 -3] | -42.6 [-49.9 -35.4] | 28.2 [20.9 35.4] | 35.4 | 1.2 [1.2 1.4] | 14.4 [14.4 16.9] |
| | WW | 43.6 (14) | 62.1 (11.4) | -18.5 (17.8) [-22.9 -14.1] | -53.3 [-60.9 -45.8] | 16.3 [8.7 23.9] | 34.8 | 1.6 [1.6 1.9] | 20.8 [20.8 24.7] |
| DUR_REM (% TST) | WSA | 15.8 (10.6) | 14.4 (5.6) | 1.4 (11.8) [-1.4 4.2] | -21.8 [-26.6 -17.1] | 24.6 [19.9 29.3] | 23.2 | 1.1 [1.1 1.3] | 32.5 [32.5 38.8] |

**Table S4. Between-device associations between raw sleep measures of single-night and 7day aggregates.**

Repeated-measures correlations (rmcorr) were computed between all pairs of measures across devices for the single-night estimate. All values were statistically significant (q < 0.001) after FDR correction. For 7-day aggregates, Pearson's correlation was used for the 7-day aggregates.

| Measure 1 | Measure 2 | Single-night [rmcorr] | 7-day aggregates [Pearson's] |
|---|---|---|---|
| mWT | wWT | 0.58 | 0.83 |
| xWT | mWT | 0.5 | 0.75 |
| xTST | mTST | 0.46 | 0.59 |
| xWT | wWT | 0.45 | 0.71 |
| mTST | wTST | 0.45 | 0.53 |
| mRPT | wRPT | 0.43 | 0.54 |
| mBT | wBT | 0.42 | 0.82 |
| mSleep_Score | wSleep_Score | 0.41 | 0.44 |
| xTST | wTST | 0.4 | 0.73 |
| xBT | mBT | 0.38 | 0.56 |
| mDUR_LS | wDUR_LS | 0.3 | 0.36 |
| mNAW | wNAW | 0.29 | 0.36 |
| xBT | wBT | 0.28 | 0.56 |
| sRPT | mRPT | 0.25 | 0.74 |
| sWT | mWT | 0.25 | 0.64 |
| sTST | mTST | 0.22 | 0.36 |
| sRPT | wRPT | 0.21 | 0.5 |
| sWT | xWT | 0.21 | 0.48 |
| mDUR_DS | wDUR_DS | 0.2 | 0.25 |
| sBT | wBT | 0.18 | 0.51 |
| mWASO | wWASO | 0.18 | 0.31 |
| sTST | wTST | 0.17 | 0.28 |
| sBT | mBT | 0.16 | 0.6 |
| xNAW | wNAW | 0.16 | 0.32 |
| sWT | wWT | 0.15 | 0.64 |
| sBT | xBT | 0.15 | 0.48 |
| sNAW | wNAW | 0.14 | 0.41 |
| sSPT | xSPT | 0.12 | 0.52 |
| mMST | wMST | 0.12 | 0.39 |

**Table S5: Single night reliability, ICC(1,1) for all sleep measures**

| Sleep Aspect | Sleep measure | Sleep diary (s) | Axivity (x) | Withing sleep analyser (m) | Withings Watch (w) |
|---|---|---|---|---|---|
| Timing | Bed Time (BT) | 0.44 [0.27 0.68] | 0.13 [0.04 0.33] | 0.4 [0.25 0.61] | 0.31 [0.17 0.52] |
| | Wake Time (WT) | 0.5 [0.34 0.71] | 0.36 [0.21 0.61] | 0.46 [0.3 0.67] | 0.2 [0.09 0.39] |
| | Mid Sleep Time (MST) | 0.59 [0.42 0.79] | 0.0 [0.0 0.09] | 0.1 [0.03 0.26] | 0.0 [0.0 0.09] |
| Duration | Total Sleep Time (TST) | 0.46 [0.3 0.67] | 0.46 [0.29 0.7] | 0.39 [0.24 0.61] | 0.3 [0.17 0.51] |
| | Sleep Period Time (SPT) | 0.47 [0.32 0.68] | 0.36 [0.2 0.6] | NA | NA |
| | Recording Period Time (RPT) | 0.59 [0.43 0.77] | NA | 0.61 [0.45 0.78] | 0.27 [0.14 0.48] |
| | TotalSleep | 0.41 [0.26 0.63] | 0.48 [0.31 0.71] | NA | NA |
| Continuity | Wake After Sleep Onset (WASO) | 0.23 [0.11 0.43] | 0.26 [0.13 0.5] | 0.54 [0.37 0.73] | 0.12 [0.04 0.29] |
| | Number of Awakenings (NAW) | 0.58 [0.42 0.77] | 0.39 [0.23 0.63] | 0.54 [0.38 0.73] | 0.28 [0.15 0.49] |
| | Out of Bed events (OOB) | NA | NA | 0.31 [0.17 0.52] | NA |
| | Sleep Efficiency(SEFF) | 0.19 [0.08 0.41] | 0.28 [0.15 0.53] | 0.5 [0.33 0.7] | 0.14 [0.06 0.32] |
| | Sleep onset latency (SOL) | 0.01 [0.0 0.14] | NA | 0.34 [0.2 0.56] | 0.02 [0.0 0.12] |
| Nap / Daytime sleep | Number of Naps (NNAP) | 0.13 [0.05 0.31] | 0.13 [0.05 0.3] | 0.0 [0.0 0.07] | NA |
| | Nap Duration (DUR_NAP) | 0.26 [0.14 0.47] | 0.13 [0.05 0.3] | 0.0 [0.0 0.07] | NA |
| Subj. Quality | Rest Upon Awakening (RuA) | 0.29 [0.12 0.63] | NA | NA | NA |
| | Quality of Sleep (QoS) | 0.42 [0.23 0.69] | NA | NA | NA |
| | Duration of Deep Sleep (DUR_DS) | NA | NA | 0.67 [0.52 0.82] | 0.52 [0.36 0.72] |
| | Duration of Light Sleep (DUR_LS) | NA | NA | 0.46 [0.3 0.67] | 0.38 [0.24 0.6] |
| | Duration of REM Sleep (DUR_REM) | NA | NA | 0.52 [0.35 0.72] | NA |
| | Number of REM bouts (NbREM) | NA | NA | 0.37 [0.23 0.59] | NA |
| | Sleep_Score | NA | NA | 0.33 [0.19 0.55] | 0.37 [0.21 0.63] |
| | Apnea hypopnea index (AHI) | NA | NA | 0.66 [0.5 0.82] | NA |

**Table S6: Fourteen night aggregated reliability for all sleep measures**

| Sleep Aspect | Sleep measure | Sleep diary (s) | Axivity (x) | Withing sleep analyser (m) | Withings Watch (w) |
|---|---|---|---|---|---|
| Timing | Bed Time (BT) | 0.92 [0.84 0.97] | 0.68 [0.37 0.87] | 0.9 [0.82 0.96] | 0.86 [0.74 0.94] |
| | Wake Time (WT) | 0.93 [0.88 0.97] | 0.89 [0.79 0.96] | 0.92 [0.86 0.97] | 0.77 [0.58 0.9] |
| | Mid Sleep Time (MST) | 0.95 [0.91 0.98] | 0.0 [0.0 0.58] | 0.62 [0.3 0.83] | 0.06 [0.0 0.58] |
| Duration | Total Sleep Time (TST) | 0.92 [0.86 0.97] | 0.92 [0.85 0.97] | 0.9 [0.82 0.96] | 0.86 [0.74 0.94] |
| | Sleep Period Time (SPT) | 0.93 [0.87 0.97] | 0.89 [0.78 0.95] | NA | NA |
| | Recording Period Time (RPT) | 0.95 [0.91 0.98] | NA | 0.96 [0.92 0.98] | 0.84 [0.7 0.93] |
| | TotalSleep | 0.91 [0.83 0.96] | 0.93 [0.86 0.97] | NA | NA |
| Continuity | Wake After Sleep Onset (WASO) | 0.8 [0.63 0.91] | 0.83 [0.68 0.93] | 0.94 [0.89 0.97] | 0.66 [0.37 0.85] |
| | Number of Awakenings (NAW) | 0.95 [0.91 0.98] | 0.9 [0.81 0.96] | 0.94 [0.9 0.97] | 0.84 [0.71 0.93] |
| | Out of Bed events (OOB) | NA | NA | 0.86 [0.74 0.94] | NA |
| | Sleep Efficiency(SEFF) | 0.76 [0.55 0.91] | 0.85 [0.71 0.94] | 0.93 [0.87 0.97] | 0.7 [0.47 0.87] |
| | Sleep onset latency (SOL) | 0.17 [0.0 0.7] | NA | 0.88 [0.78 0.95] | 0.23 [0.0 0.66] |
| Nap / Daytime sleep | Number of Naps (NNAP) | 0.68 [0.42 0.86] | 0.68 [0.42 0.86] | 0.0 [0.0 0.51] | NA |
| | Nap Duration (DUR_NAP) | 0.83 [0.7 0.93] | 0.68 [0.42 0.86] | 0.0 [0.0 0.51] | NA |
| Subj. Quality | Rest Upon Awakening (RuA) | 0.85 [0.66 0.96] | NA | NA | NA |
| | Quality of Sleep (QoS) | 0.91 [0.81 0.97] | NA | NA | NA |
| | Duration of Deep Sleep (DUR_DS) | NA | NA | 0.97 [0.94 0.98] | NA |
| | Duration of Light Sleep (DUR_LS) | NA | NA | 0.92 [0.86 0.97] | NA |
| | Duration of REM Sleep (DUR_REM) | NA | NA | 0.94 [0.88 0.97] | NA |
| | Number of REM bouts (NbREM) | NA | NA | 0.89 [0.81 0.95] | NA |
| | Sleep_Score | NA | NA | 0.87 [0.77 0.94] | 0.89 [0.79 0.96] |
| | Apnea hypopnea index (AHI) | NA | NA | 0.96 [0.93 0.98] | NA |

**Table S7. Between-device associations between PCA derived sleep aspects of single-night and 7-day aggregates.**

Repeated-measures correlations (rmcorr) were computed between all pairs of measures across devices for the single-night PCA scores. All values were statistically significant ($q < 0.001$) after FDR correction. For 7-day aggregates, Pearson's correlation was used. The table lists associations as $|r|$. The * indicates PCA derived sleep aspects that reach ICC ≥ 0.7 in 7 nights.

| Measure 1 | Measure 2 | Single-night [rmcorr] | 7-day aggregates [Pearson's] |
|---|---|---|---|
| **Raw PCA derived sleep aspects** | | | |
| mDuration | wDuration | 0.45 | 0.52 |
| xDuration | mDuration | 0.42 | 0.61 |
| xDuration | wDuration | 0.39 | 0.68 |
| xTiming* | mTiming* | 0.36 | 0.6 |
| mTiming* | wTiming* | 0.34 | 0.75 |
| xTiming* | wTiming* | 0.31 | 0.58 |
| sDuration | mDuration | 0.28 | 0.67 |
| sDuration | wDuration | 0.23 | 0.48 |
| mNREM-AHI | wNREM | 0.23 | 0.31 |
| sDuration | xDuration | 0.19 | 0.42 |
| sTiming | xTiming* | 0.18 | 0.41 |
| sQuality | wContinuity* | 0.16 | 0.25 |
| xContinuity | mContinuity | 0.12 | 0.32 |
| xContinuity | wContinuity* | 0.11 | 0.3 |
| sTiming | wTiming* | 0.11 | 0.55 |
| **Clean PCA derived sleep aspects** | | | |
| xDuration | mDuration | 0.42 | 0.61 |
| mDuration | wDuration | 0.38 | 0.4 |
| xDuration | wDuration | 0.35 | 0.59 |
| sDuration | mDuration | 0.28 | 0.66 |
| sDuration | wDuration | 0.22 | 0.42 |
| sDuration | xDuration | 0.19 | 0.41 |
| mEfficiency | wDuration | 0.14 | 0.25 |
| sTiming | mEfficiency | 0.13 | 0.4 |
| mNREM-AHI | wContinuity | 0.13 | 0.29 |
| xContinuity | mContinuity | 0.12 | 0.3 |

**Table S8. Discriminative performance of the devices – Raw sleep measures.**

Values shown are the mean followed by the (standard deviation) of 5-fold cross validation and [min max]

| Device | Area under the curve (AUC) | Average precision (AP) |
|---|---|---|
| **Pooled** | 0.70 (0.09)<br>[0.57, 0.77] | 0.49 (0.20)<br>[0.18, 0.68] |
| **Sleep Diary** | 0.64 (0.10)<br>[0.54, 0.79] | 0.45 (0.23)<br>[0.08, 0.69] |
| **Axivity** | 0.59 (0.16)<br>[0.34, 0.76] | 0.46 (0.24)<br>[0.07, 0.64] |
| **WSA** | 0.44 (0.08)<br>[0.33, 0.52] | 0.26 (0.15)<br>[0.07, 0.46] |
| **WW** | 0.50 (0.12)<br>[0.36, 0.70] | 0.33 (0.22)<br>[0.08, 0.63] |

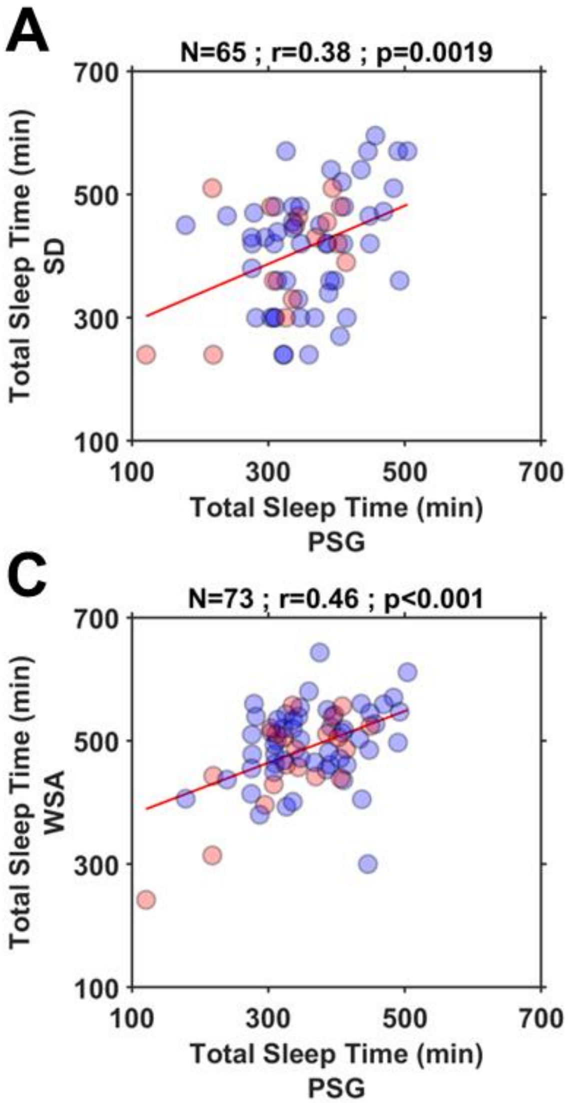


**Figure S1. Associations between the total sleep time (TST) estimated by the Devices vs Polysomnography (PSG). A. Sleep Diary (SD), B. Axivity (AX), C. Withings sleep analyser (WSA) and D. Withings Watch (WW).** The data points in red depicts people living with Dementia, blue depicts controls. All device measures are automatically estimated by the respective proprietary algorithm. The top of each of the plots shows the number of participants, the Pearson's correlation and significance value of the association between the devices.

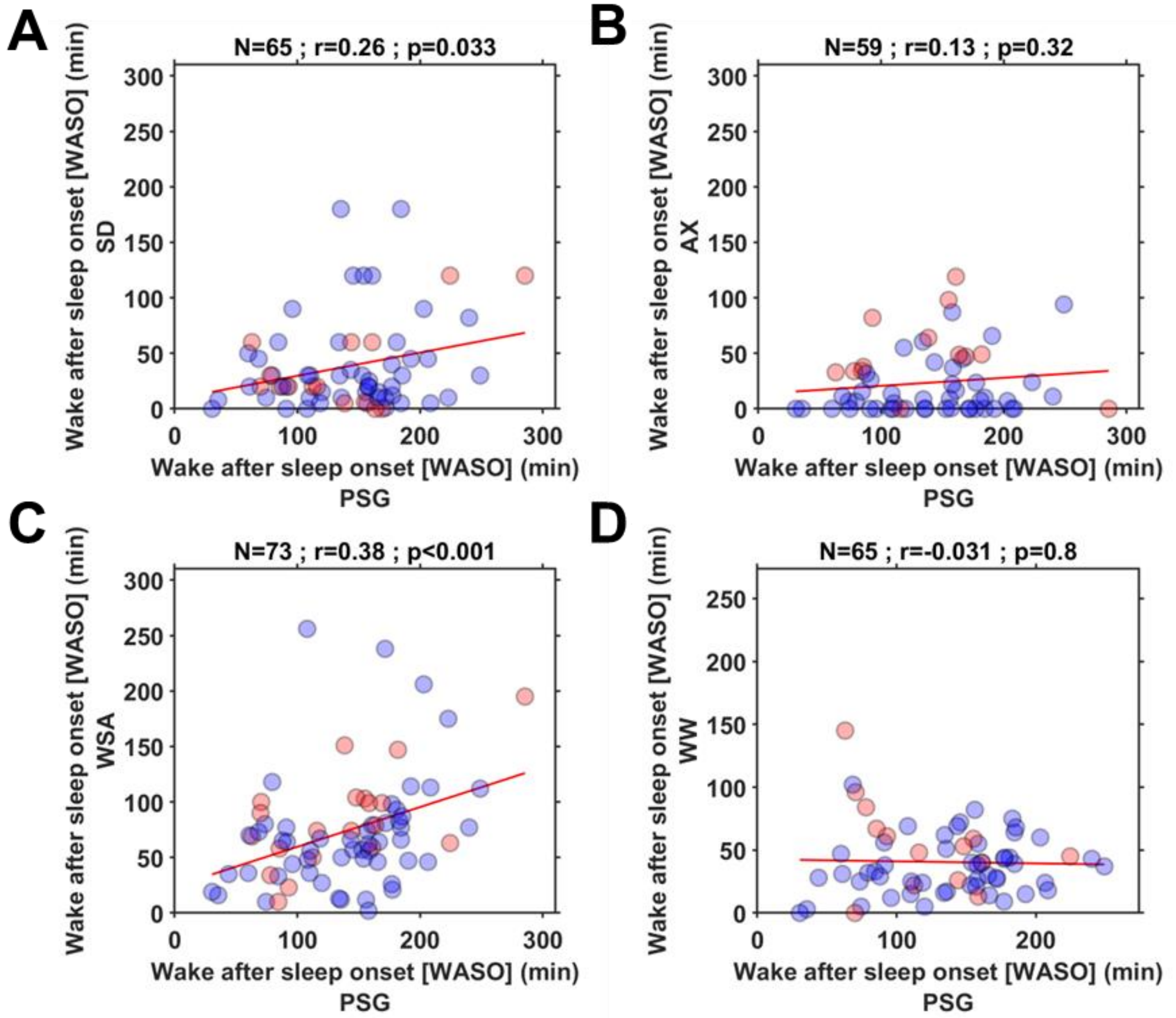


**Figure S2. Associations between the wake after sleep onset (WASO) estimated by the Devices vs Polysomnography (PSG). A. Sleep Diary (SD), B. Axivity (AX), C. Withings sleep analyser (WSA) and D. Withings Watch (WW).** The data points in red depicts people living with Dementia, blue depicts controls. All device measures are automatically estimated by the respective proprietary algorithm. The top of each of the plots shows the number of participants, the Pearson's correlation and significance value of the association between the devices.

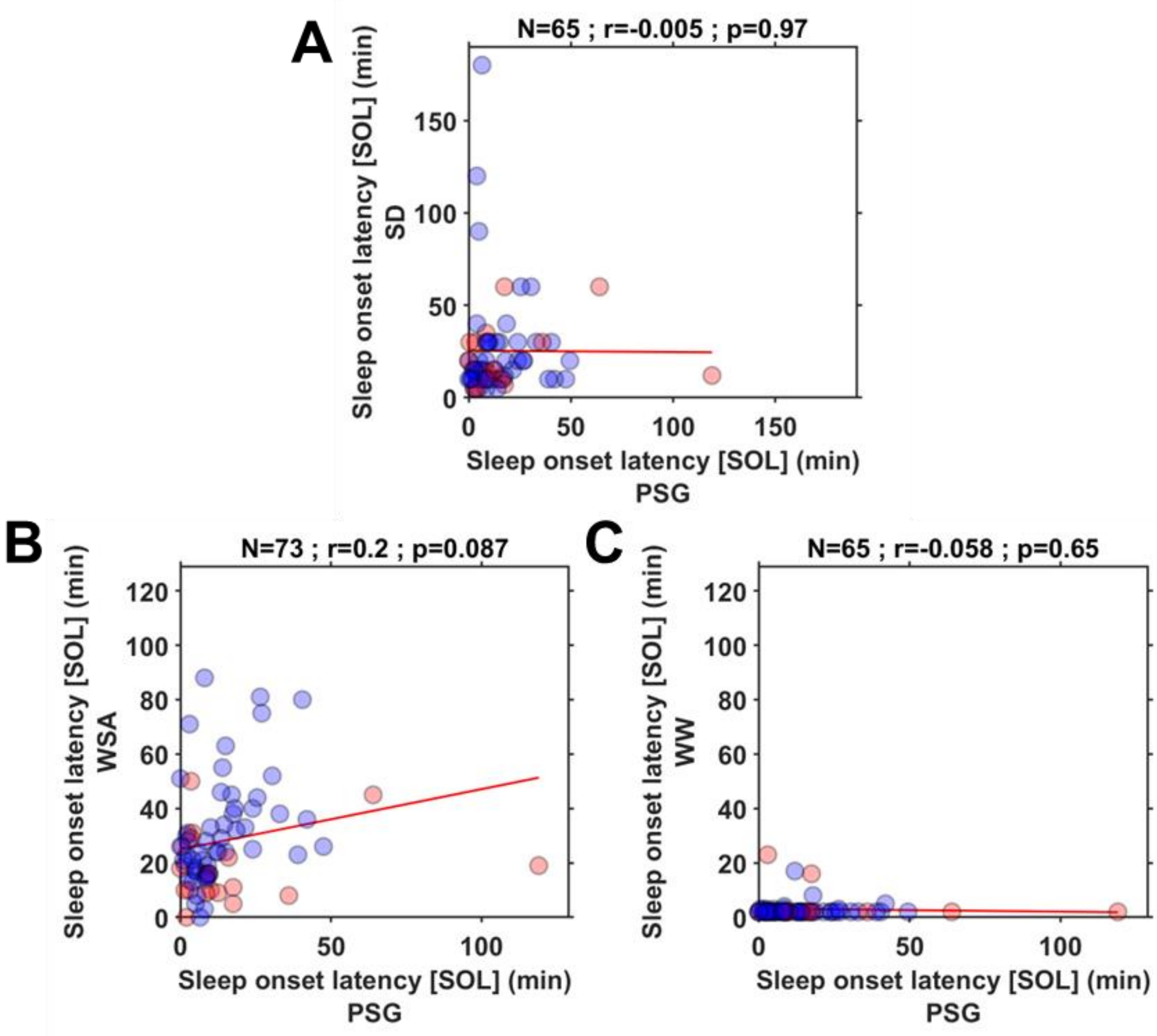


**Figure S3. Associations between the sleep onset latency (SOL) estimated by the Devices vs Polysomnography (PSG). A. Sleep Diary (SD), B. Withings sleep analyser (WSA) and D. Withings Watch (WW).** The data points in red depicts people living with Dementia, blue depicts controls. All device measures are automatically estimated by the respective proprietary algorithm. The top of each of the plots shows the number of participants, the Pearson's correlation and significance value of the association between the devices.

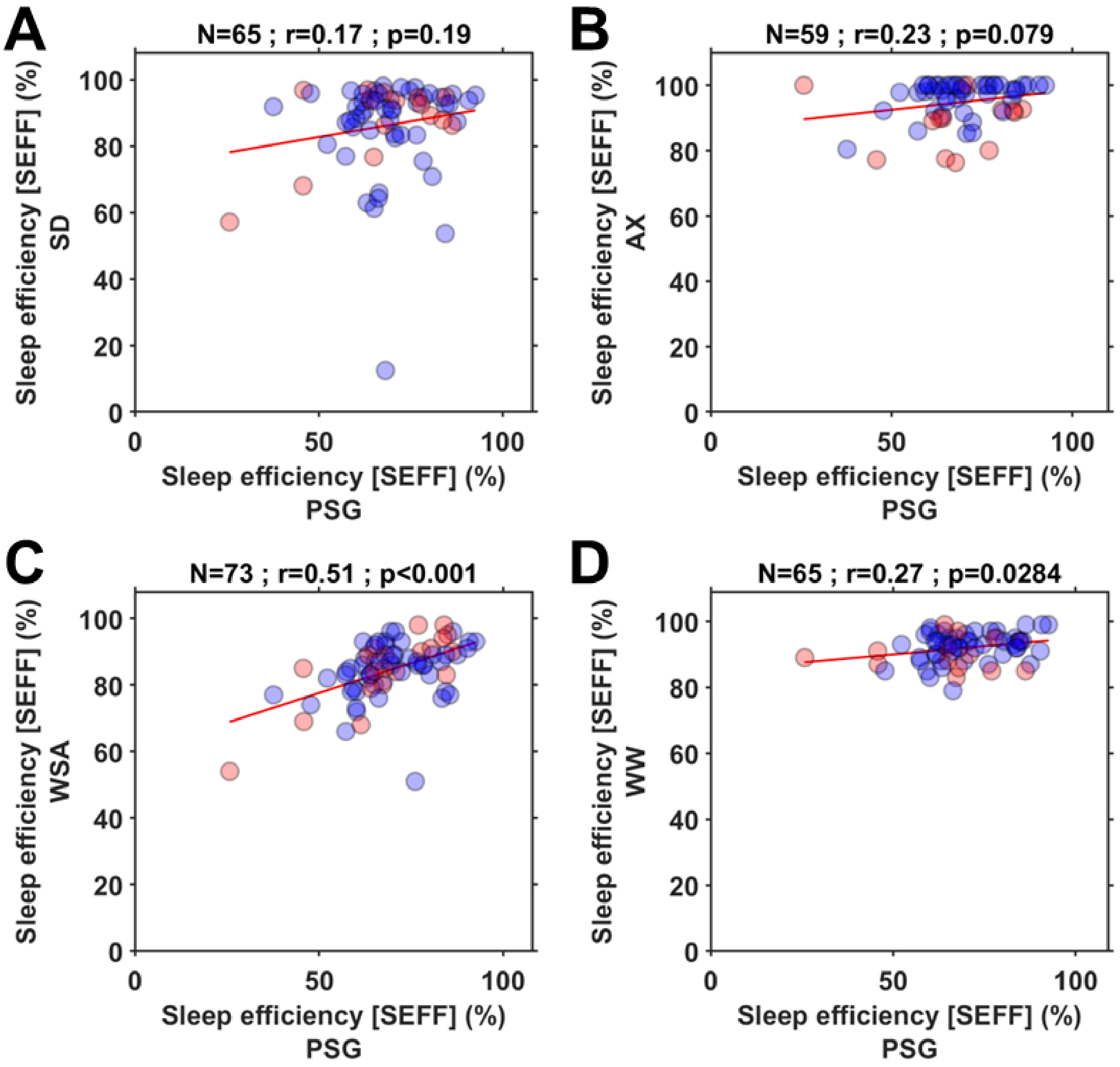


**Figure S4. Associations between the sleep efficiency (SEFF) estimated by the Devices vs Polysomnography (PSG). A. Sleep Diary (SD), B. Withings sleep analyser (WSA) and D. Withings Watch (WW).** The data points in red depicts people living with Dementia, blue depicts controls. All device measures are automatically estimated by the respective proprietary algorithm. The top of each of the plots shows the number of participants, the Pearson's correlation and significance value of the association between the devices.

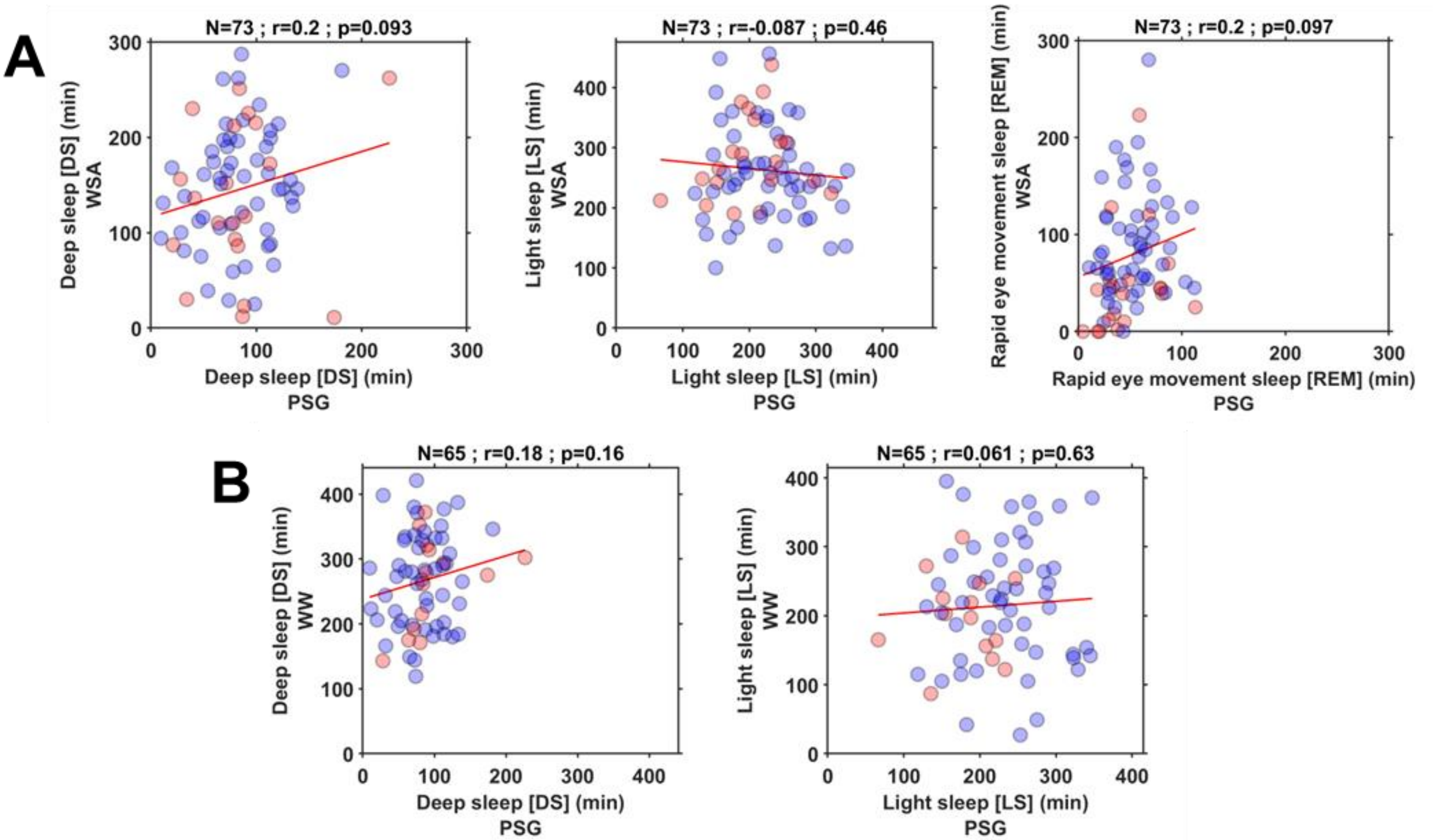


**Figure S5. Associations between the sleep stage durations of Withings Devices and Polysomnography. A. Withings sleep analyser (WSA) B. Withings Watch (WW).** The data points in red depicts people living with Dementia, blue depicts controls. All device measures are automatically estimated by the respective proprietary algorithm. The top of each of the plots shows the number of participants, the Pearson's correlation and significance value of the association between the devices

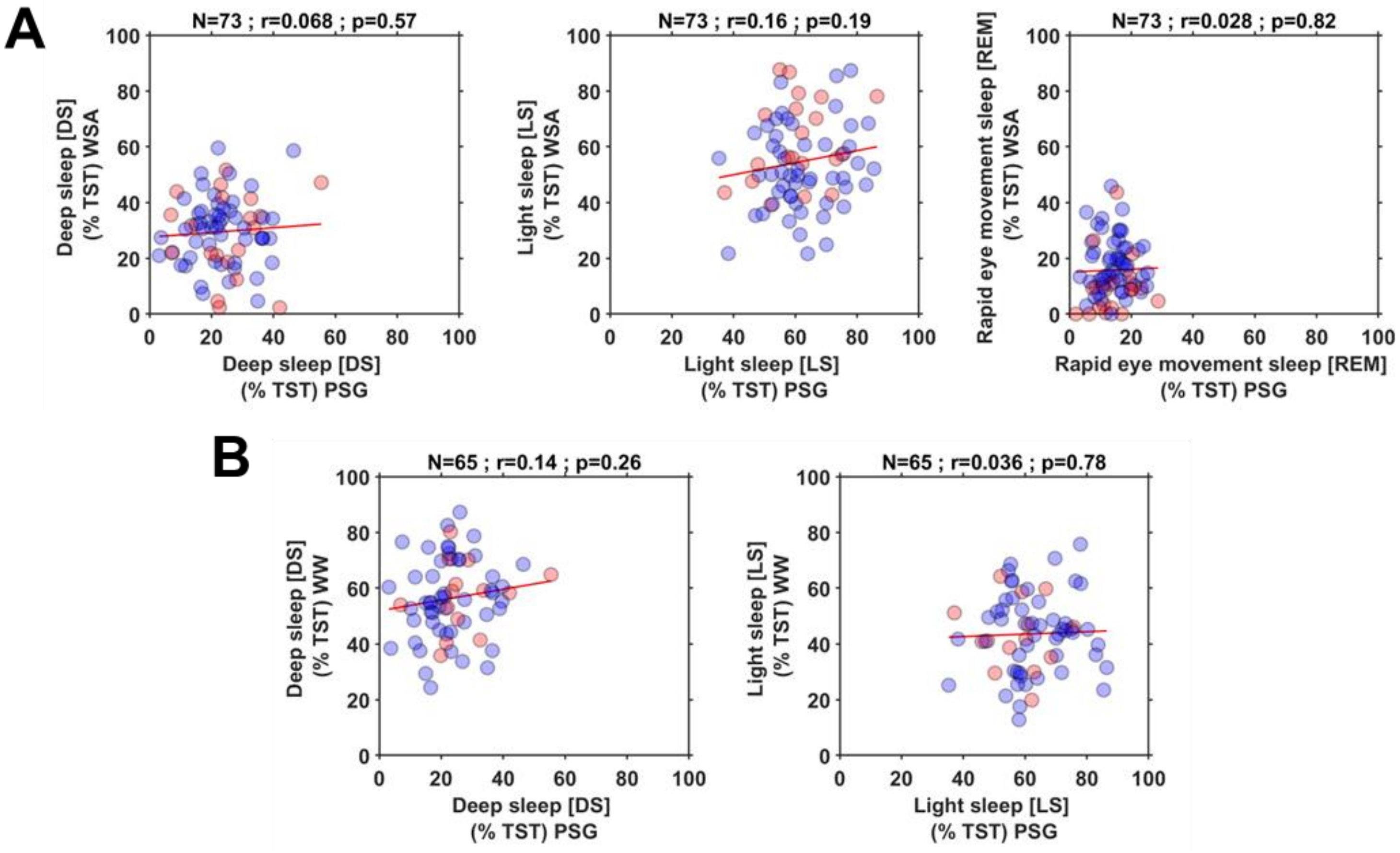


**Figure S6. Associations between the sleep stage durations expressed as percentage of total sleep time (TST) of Withings Devices and Polysomnography. A. Withings sleep analyser (WSA) B. Withings Watch (WW).** The data points in red depicts people living with Dementia, blue depicts controls. All device measures are automatically estimated by the respective proprietary algorithm. The top of each of the plots shows the number of participants, the Pearson's correlation and significance value of the association between the devices

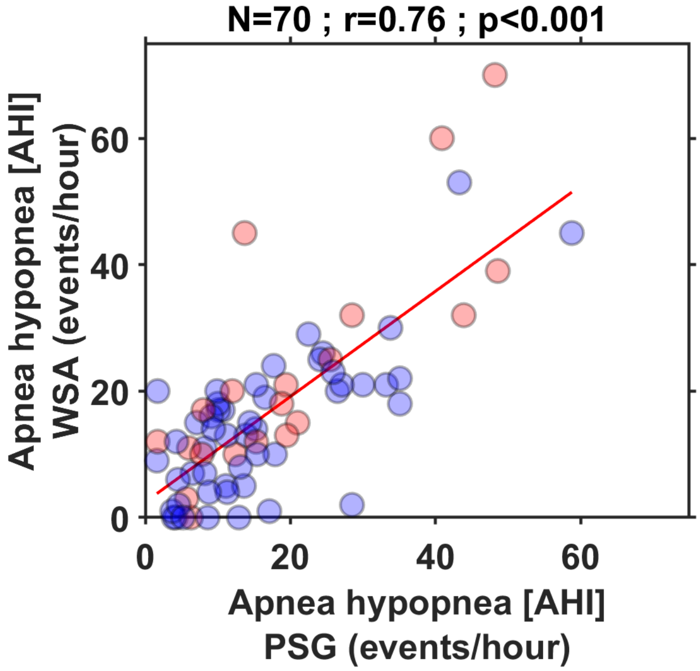


**Figure S7. Associations between the sleep measures of Withings sleep analyser apnea-hypopnea index (AHI) and Polysomnography.** The data points in red depicts people living with Dementia, blue depicts controls. All device measures are automatically estimated by the respective proprietary algorithm. The top of each of the plots shows the number of participants, the Pearson's correlation and significance value of the association between the devices.

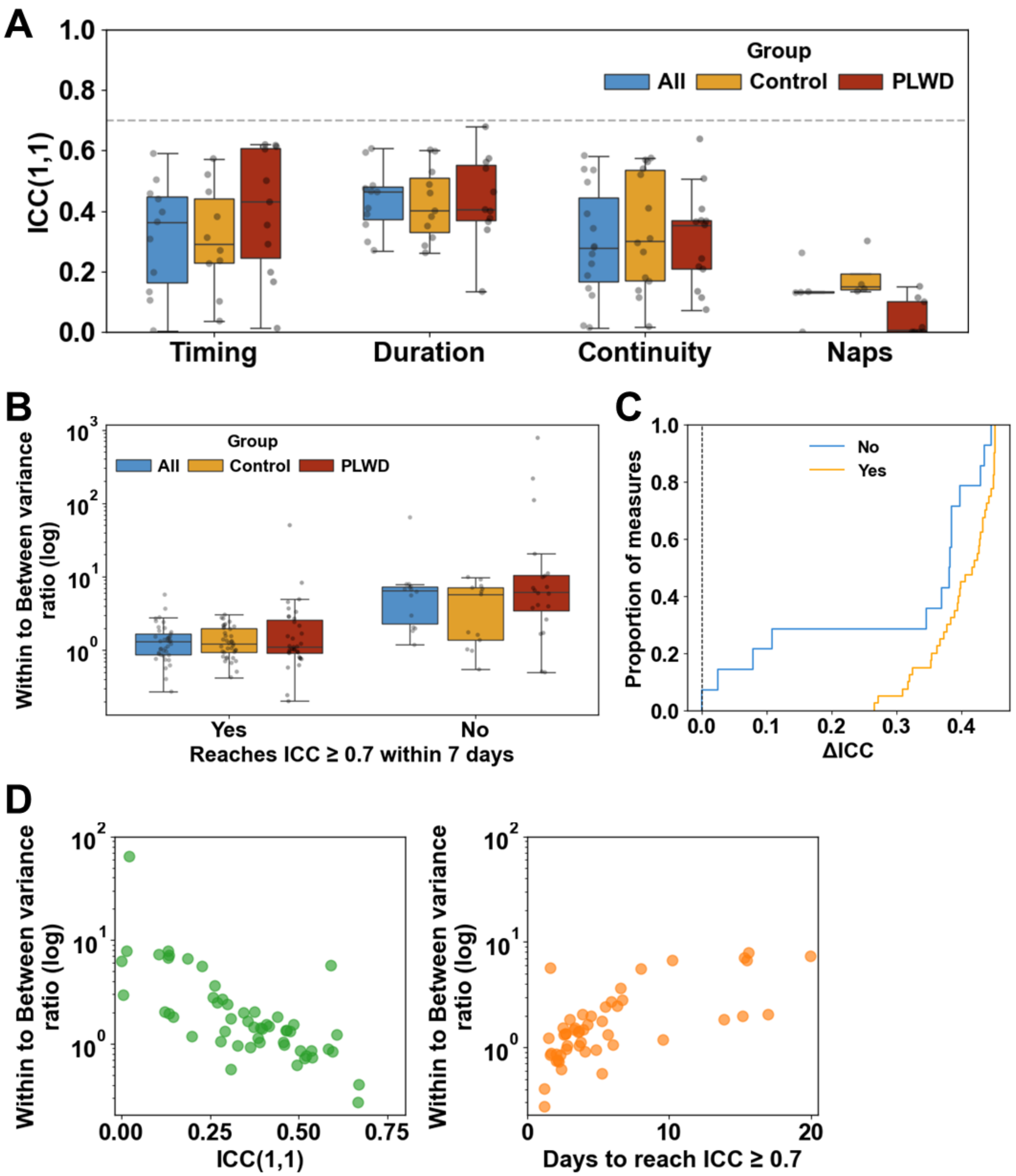


**Figure S8. Reliability characteristics of sleep measures. A.** Single-night reliability (ICC(1,1)) across sleep aspects (Timing: Get to bed time (BT), Wake time/Get out of bed time (WT), Mid sleep time (MST); Duration: Total sleep time (TST), Sleep period time (SPT), Recording period time (RPT), 24 hour Total sleep time (TotalSleep); Continuity: Wake after sleep onset (WASO), Sleep efficiency (SEFF), Number of awakenings (NAW), Sleep onset latency (SOL); Naps: Duration of naps (DUR_NAP); Number of naps (NNAP)). **B.** Within to between participant variance ratio and **C.** Cumulative distribution of ΔICC, for the measures that reach (Yes) and those that fail (No) to reach acceptable reliability (ICC ≥ 0.7) in 7 days of aggregation. **D.** Relationship between within to between participant variance ratio vs ICC and the number of days of aggregation required to reach acceptable reliability (ICC ≥ 0.7).

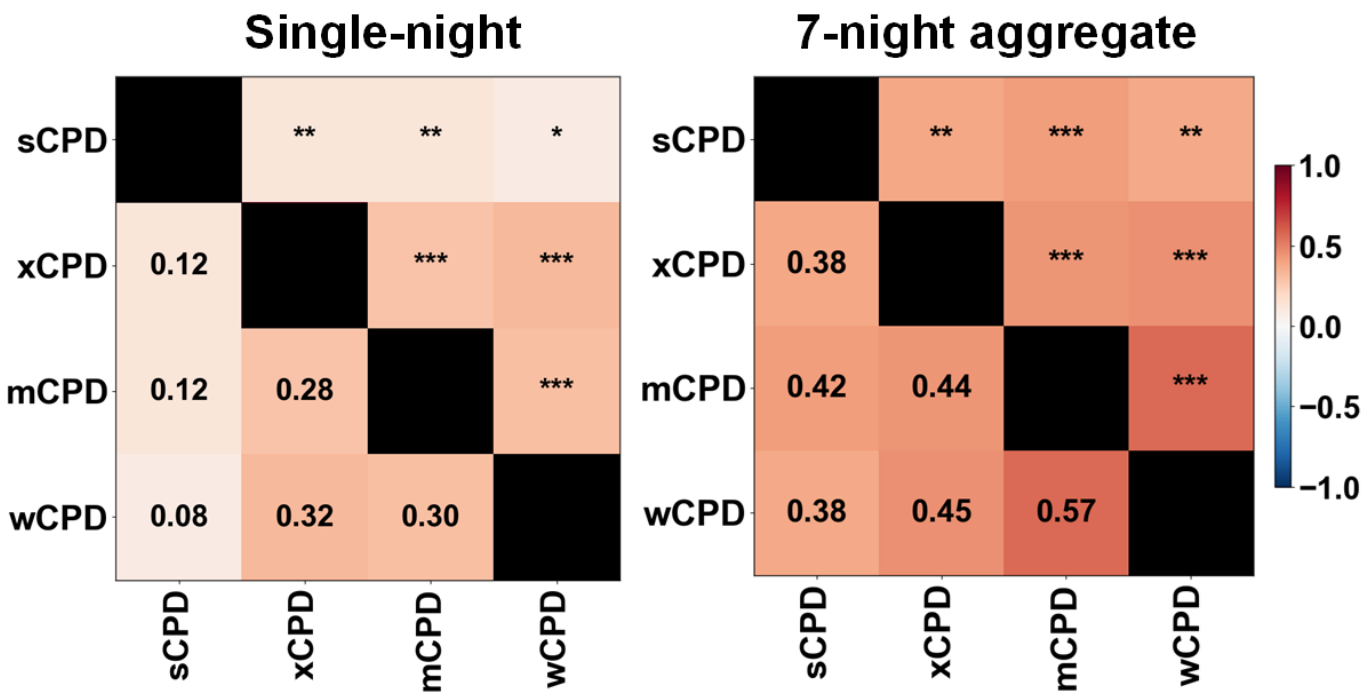


**Figure S9. Between-device associations for Composite Phase Deviation (CPD).** Heatmaps show between-device correlations for single-night CPD (left) and CPD aggregated across 7 nights (right). CPD was estimated using participant-specific mean midsleep time (MST) calculated from all available nights. Repeated-measures correlations (rmcorr) were computed for the single-night CPD. For 7-day aggregates, Pearson's correlation was used. Asterisks denote significance levels (* q/p<0.05, ** q/p <0.01, *** q/p <0.001).

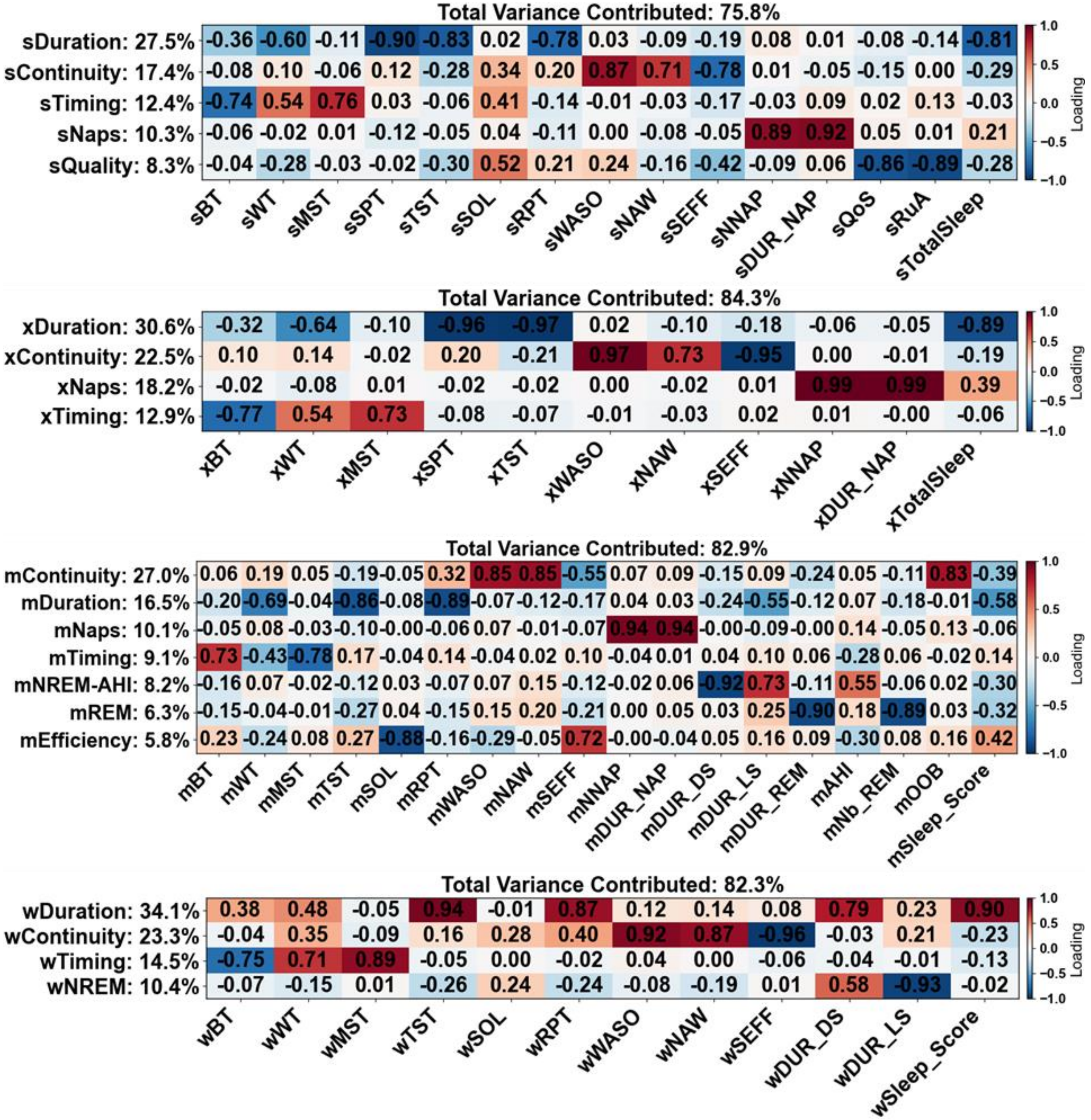


**Figure S10. Principal component analysis (PCA) of all the raw sleep measures.** The device components are marked with s – Sleep Diary; x – Axivity; m – Withings sleep analyser (WSA); w – Withings Watch (WW). Identified component labels and the percentage of variance explained are shown on the left of the heatmap of the standardized loadings, and total variance in the device explained components with eigen values over 1 is indicated above each panel.

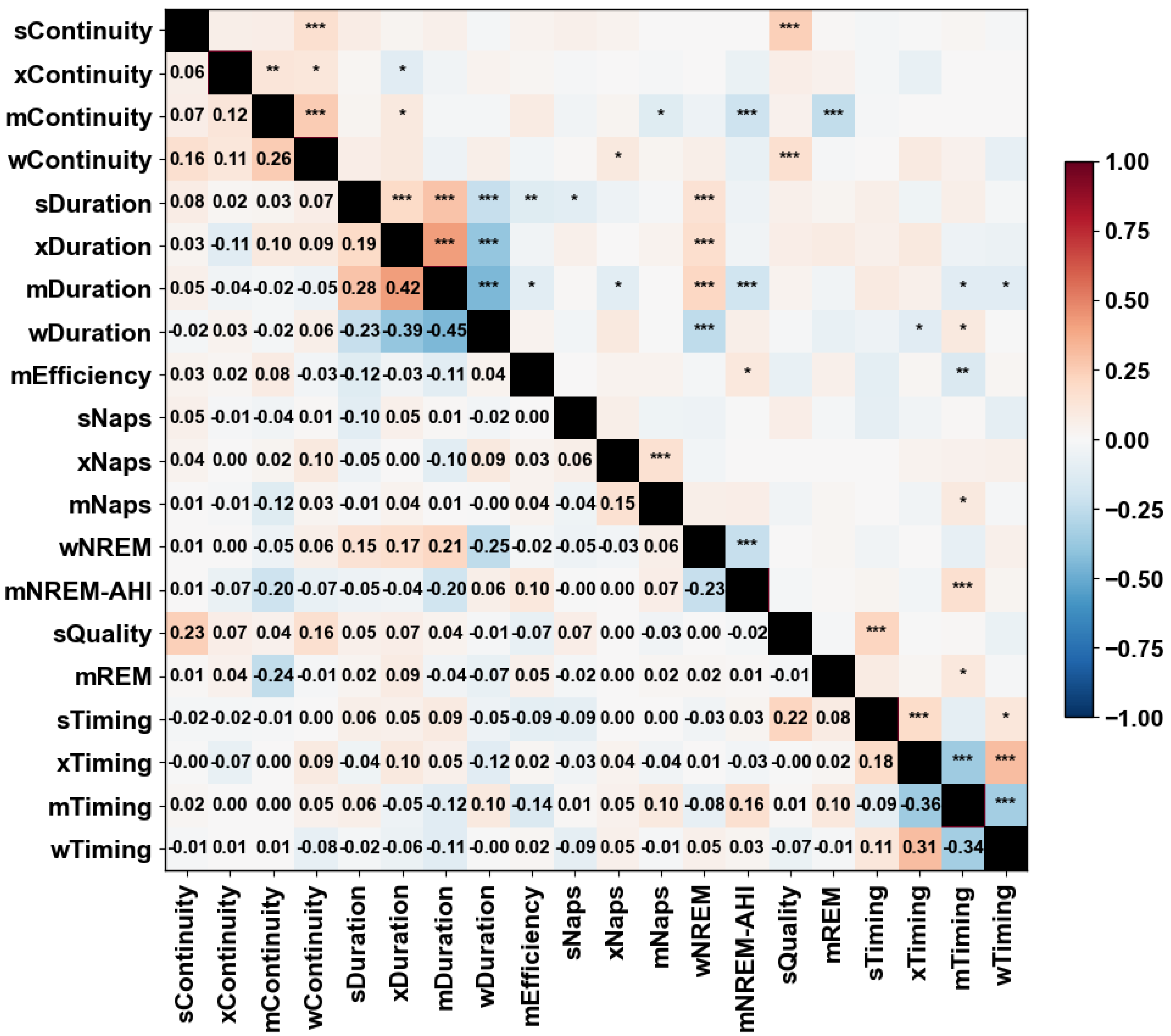

**Figure S11. Heatmap of the single-night associations between PCA-derived sleep aspects (PSAs) identified across devices.** All the raw sleep measures were used in the analysis. Correlation estimated: repeated-measures correlation; multiple testing correction was performed using the Benjamini-Hochberg false discovery rate and adjusted q-values are provided; asterisks denote significance levels (* q<0.05, ** q<0.01, *** q<0.001).

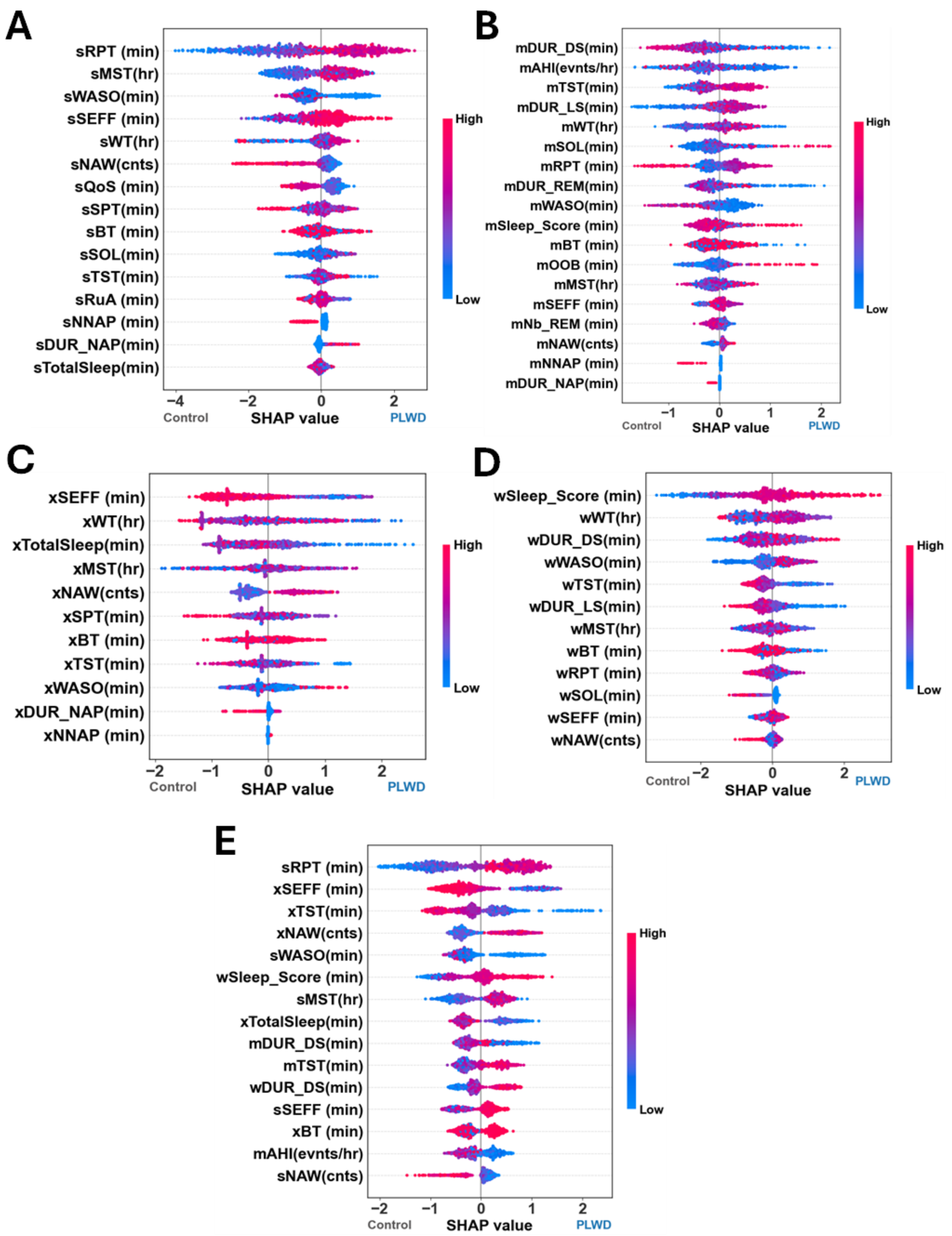


**Figure S12. SHapley Additive exPlanations (SHAP) values depicting the impact of each measure on the Control vs PLWD discrimination performance. A.** Sleep Diary (s), **B.** Withings sleep analyser (m), **C.** Axivity (x), **D.** Withings watch (w) and **E.** Measures pooled across all devices. The sleep measures in each plot are ordered vertically based on mean absolute SHAP values. Each data point comes from nights in the held-out data from the five-fold participant-wise cross-validation. The horizontal dispersion of each point denotes the direction and magnitude of the sleep measures contribution to the model prediction (negative - Control, positive - PLWD). Red indicates high sleep measure values while blue indicates low values. For example, a red data point with a positive SHAP value indicates that higher values of that sleep measure contribute positively to the probability of classification of PLWD, whereas a blue data point with a negative SHAP value indicates that lower values contribute towards classification as Control.

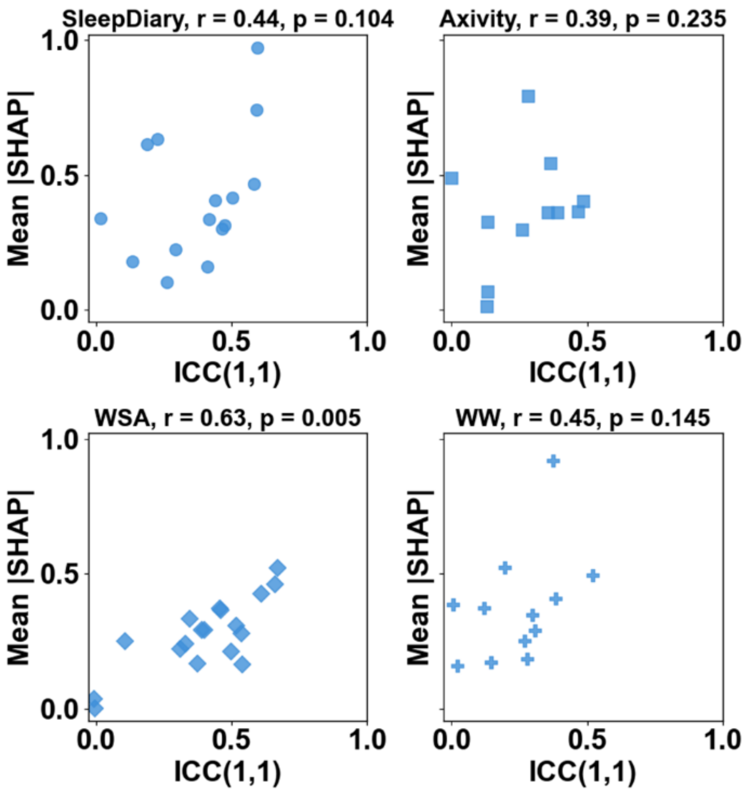


**Figure S13. Relationship between single night reliability (ICC(1,1)) and Mean SHapley Additive exPlanations (SHAP) values.** The device name and the spearman correlation is provided at the top of the scatter plots. Total number of measures for each device are: Sleep diary, n=15; Axivity, n=11; Withings sleep analyser (WSA), n=18; Withings watch (WW), n=12.
